# Supporting the revision of the health benefits package in Uganda: a constrained optimisation approach

**DOI:** 10.1101/2022.05.04.22274650

**Authors:** Sakshi Mohan, Simon Walker, Freddie Sengooba, Elizabeth Ekirapa Kiracho, Chrispus Mayora, Aloysius Ssennyonjo, Candia Tom Aliti, Paul Revill

## Abstract

This study demonstrates how the linear constrained optimization approach can be used to design a health benefits package (HBP) which maximises the net disability adjusted life years (DALYs) averted given the health system constraints faced by a country, and how the approach can help assess the marginal value of relaxing health system constraints. In the analysis performed for Uganda, 58 interventions were included in the HBP in the base scenario, resulting in a total of 49.9 million net DALYs averted. When task shifting of pharmacists’ and nutrition officers’ tasks to nurses is allowed, 68 interventions were included in the HBP resulting in a total of 53.8 million net DALYs averted (a 7.8% increase). Further, investing only $39 towards hiring additional nutrition officers’ time could avert one net DALY; this increased to $55, $56, and $123 for nurses, pharmacists and doctors respectively, and $971 for expanding the consumable budget.

## Introduction

All public health systems are faced with the crucial question of how best to allocate their limited resources in order to maximise the benefits they produce. This question is particularly pressing in low- and middle-income countries (LMICs) where the resource constraints, financial and other, mean that many health services cannot be provided to all those in need(1). It is widely recognized that there is a considerable gap between the aspirational health plans of LMICs and actually available resources(2,3). Health Benefits Packages offer a potential solution to the implicit sub-optimal rationing of resources(4) which occurs as a result of this mismatch. By changing from ad hoc or implicit priority setting and rationing of services, to systematic, evidence-based and transparent priority setting based on cost-effectiveness analysis (CEA), countries can substantially improve health outcomes, improve access to important high-quality services and achieve national and global sustainable development goals (SDG) targets(5).

In the context of HBPs, CEA is concerned with maximizing the population health benefits (considering other priorities such as equity) obtained from the services in the package given the resources available. This can be characterized in the form of a constrained optimization problem (6). The approach has previously been applied in studies optimizing the distribution of a specific intervention among the eligible population(7–9), studies optimizing the choice of interventions within a single disease area or programme (10,11), and theoretical analyses (12–17). The approach has also been discussed in broader methods guidelines(5,18–21). Among the empirical studies, those which took a more comprehensive view of the health sector rather than a specific disease or programme only applied an ‘overall’ financial constraint facing a health system reflecting the financial cost of all resource inputs(22,23). While such an approach may be suitable over the long run over which all resources are potentially flexible, in the short run, there are multiple constraints on care both financial and non-financial(24–26). There is a need, therefore, for empirical research to demonstrate how other resource constraints can be captured in conjunction with the public health budget constraint to arrive at an optimal HBP.

This study demonstrates the use of a linear constrained optimization approach to develop a health benefits package which maximises the net health impact (or net disability adjusted life years, DALYs, lost averted) given the financial and physical resource constraints of Uganda’s public health sector. Recognising the “human resources for health crisis” (27) in the region, this study focuses on the size and composition of the workforce in Uganda to capture physical resource constraints. However, the analytical framework offers the flexibility to include other health system constraints and can be applied to answer some of the most pressing resource allocation decisions facing ministries of health, for example - which interventions represent “best buys” within the health system; where investments in health systems strengthening should be made and how much the government can afford to pay for health systems strengthening; implications of donor funding conditionalities; and the impact of task shifting among health workers.

## Methods

### Overview

A constrained optimization approach was used to identify the optimal list of services to be included in Uganda’s HBP. The approach is set up as a linear programming problem (LPP) to choose the level of optimal coverage of each possible intervention in order to maximise the population health benefit while ensuring that the resources required to deliver the interventions do not exceed those currently available in Uganda. The LPP can be represented as follows:

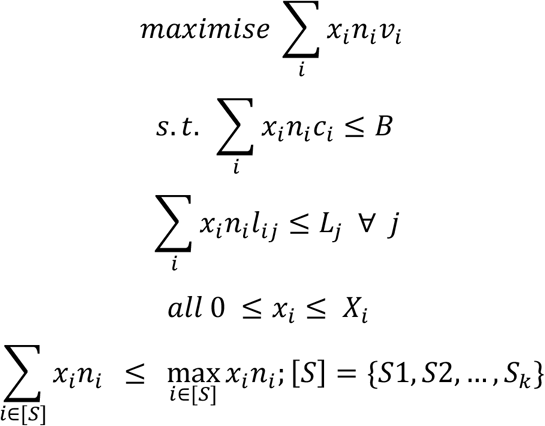

where ***x***_***i***_ = percentage of people receiving intervention i (decision variable)

***n***_***i***_ **=** number of patients eligible for intervention i

***v***_***i***_ = net DALYs averted per patient treated with intervention i

***c***_***i***_ = consumables cost of intervention i per patient

**B** = total annual budget for consumables

***l***_***ij***_ = time requirement of cadre j per patient treated with intervention i

***L***_***j***_ = total time available of cadre j

***X***_***i***_ = maximum feasible coverage of intervention i

***S***_***k***_ = list of substitutable interventions in group k

The following sub-sections describe the components of the LPP in further detail.

### Decision variables

The decision variables in our LPP are the level of coverage of various health interventions (as a percentage of people eligible for interventions) in the HBP, allowing for the possibility of complete exclusion of interventions from the package (i.e. 0% coverage). Interventions are assumed to be independent of each other, i.e. their costs and effects do not depend upon what other interventions are provided, and costs and health effects are linear in implementation, i.e. we do not reflect any economies of scale. A total of 278 interventions were considered for inclusion in the Ugandan HBP. This list was drawn from a combination of the 2015/16-2019/20 HBP for Uganda(28), as well as any cost-effective substitutes of these interventions which were found during the review of the cost-effectiveness literature. Of these 278 possible interventions, minimum data for the LPP could be obtained for 128 interventions, which were then considered for inclusion in the HBP. These interventions represent the majority of the disease burden in Uganda(29).

### Objective function

The objective is to *maximise* the health benefit provided by the chosen set of interventions, which is measured in terms of net disability adjusted life years (DALYs). Net DALYs are measured as DALYs-averted by the intervention less DALYs averted forgone as a result of limited resources being committed to the intervention not being available for other purposes (i.e. it is the health opportunity cost, measured using a cost-effectiveness threshold (CET)). Net DALYs rather than DALYs alone are used as the objective function to incorporate the consequences of future, downstream costs. The choice of CET is crucial to this calculation. Based upon a study that estimates the marginal productivities of health systems – i.e., the health consequences associated with changes in budget - the CET for Uganda was assumed to be $161 in the base case(30).

### Constraints

The constraints represent the capacity of the health system to deliver health interventions. In our analysis, we consider the following constraints – i. *the size of the resource envelope to purchase consumables*, ii. *the size of the health workforce* sub-divided among five cadres (doctors and medical officers, nurses, pharmacists, nutrition officers, and mental health officers), iii. *the maximum feasible coverage level for each intervention* (as estimated during the design of the Uganda’s current Health Sector Development Program II, 2020/21-2024/25 (HSDP)(31)). In terms of resources, only the consumables budget and health worker constraints are captured explicitly, however, the constraints on feasible coverage are expected to reflect other supply-side constraints (medical equipment, hospital capacity) and demand-side constraints. Further constraints were installed to ensure that the sum of coverage of substitutable interventions did not exceed the size of the eligible population (see supplementary table 5 for the list of substitutable interventions).

### Data

Data for the specification of parameters for these interventions were obtained from a range of sources for the year of analysis (2020) (see Table 1). For the objective function, DALYs averted per case and full healthcare cost estimates were obtained from existing economic evaluation literature, including but not limited to the Global Health Cost-Effectiveness Analysis Registry(32) and WHO-CHOICE(33). Estimates of the size of the eligible population for each intervention were obtained using inputs into the One Health Tool (OHT)(34) used to cost the HSDP II(31). Where this information was missing, estimates were obtained from published literature and reports.

**Table 1:** Data sources for model parameters.

| **Model parameter** | **Data source** |
| --- | --- |
| Cost-effectiveness of interventions (DALYs averted per person, cost per person) | Literature search, including but not limited to WHO-CHOICE (33) and the Global Health Cost-effectiveness Registry(32) |
| Eligible population | One Health Tool (OHT)(34) used to cost HSDP II 2020/21 – 2024/25<sup>1</sup>(31) |
| Maximum feasible coverage of interventions | One Health Tool (OHT)(34) used to cost HSDP II 2020/21 – 2024/25<sup>1</sup> |
| Consumables cost | One Health Tool (OHT)(34) used to cost HSDP II 2020/21 – 2024/25<sup>1</sup> |
| Health worker time required | Workforce optimization model exercise for the Malawi Human Resources for Health Strategic Plan, 2018–2022<sup>2</sup> (35) |
| Health opportunity cost | Lomas et al. (2021) (30) |
| Consumables budget | Government of Uganda(GoU) budget records |
| Health worker capacity | MoH HR records, consultation |

<sup>1</sup> The data obtained from this source was complemented with a literature search
<sup>2</sup> The data obtained from this source was complemented with inputs from staff at the Ministry of Health, Uganda

The maximum feasible coverage level for each intervention as well as the cost of consumables required for interventions were also obtained from the OHT. Estimates of health worker time required to deliver interventions came from the Workforce optimization model exercise carried out for the Malawi Human Resources for Health Strategic Plan, 2018–2022 (35) which captured the number of minutes required to provide common health services based on time-motion observations and expert opinion. In the absence of local data, we assumed that the health worker time requirements in Uganda would be identical to those recorded in Malawi.

The size of the current health workforce (2020) and average annual patient-facing time per health worker was established through consultations with the Ministry of Health, Uganda. The consumables budget was assumed to be the same as that in 2020 as obtained from the government’s budget records. These constraints were established based only on the public healthcare sector as well as the private not-for-profit sector supported financially by the government, both of which provide free or heavily subsidized health care. Supplementary table 1 provides data on all the included parameters on the 128 candidate interventions for which evidence is currently available. All monetary figures are presented in 2019 US$.

### Extensions

#### Allowing for task shifting

The acute shortage of health workers in many sub-Saharan African countries(36) means that gaps in workforce size and composition are often filled through task-shifting, particularly to nurses(37,38). In order to capture this, we consider a scenario under which nurses are able to substitute for nutrition officers and pharmacists, implying that the nutrition officer and pharmacist time required to deliver interventions is allowed to be converted to nurse time. We refer to this scenario as the task-shifting scenario.

#### Effect of removing health system constraints

We run additional scenarios excluding the non-financial constraints, i.e. the size of the health workforce and maximum feasible coverage, from the LPP. We demonstrate the effect of these omissions on the inclusion of interventions into the HBP, its health impact as well as resource use implications.

#### Estimation of the marginal value of relaxing health system constraints

The constrained optimization approach allows for additional analyses on the effect of relaxing some of the constraints applied. In particular, we assess how much health is gained from investing an additional $1000 towards each resource, i.e. towards the consumables budget and salaries of health workers (see supplementary table 6 for salary figures used). Conversely, we also present the additional investment required in each of these health system components in order to avert a single DALY at the margin. This generates an incremental cost-effectiveness ratio (ICER) of these investments, estimated as a cost-per-DALY-averted, which can be compared to results of other cost-effectiveness studies.

## Results

Under the base scenario, the linear constrained optimization approach provides an optimal HBP consisting of 58 interventions averting 49.9 million net DALYs, which can be feasibly delivered within Uganda’s health system constraints. Table 2 provides a summary of the health impact and resource requirements of this HBP. We observe that a significant proportion of the capacity of doctors/medical officers and nurses remains underutilised under this scenario due to the limited availability of pharmacists to dispense drugs. We therefore consider a more realistic case where, upon the exhaustion of pharmacist and nutrition officer time, their tasks are taken up by nurses (task shifting scenario). This results in the exhaustion of the drug budget as well as the capacity of four out of five health worker cadres while adding 10 more interventions to the optimal HBP and increasing the number of net DALYs averted by 7.7%.

**Table 2:**
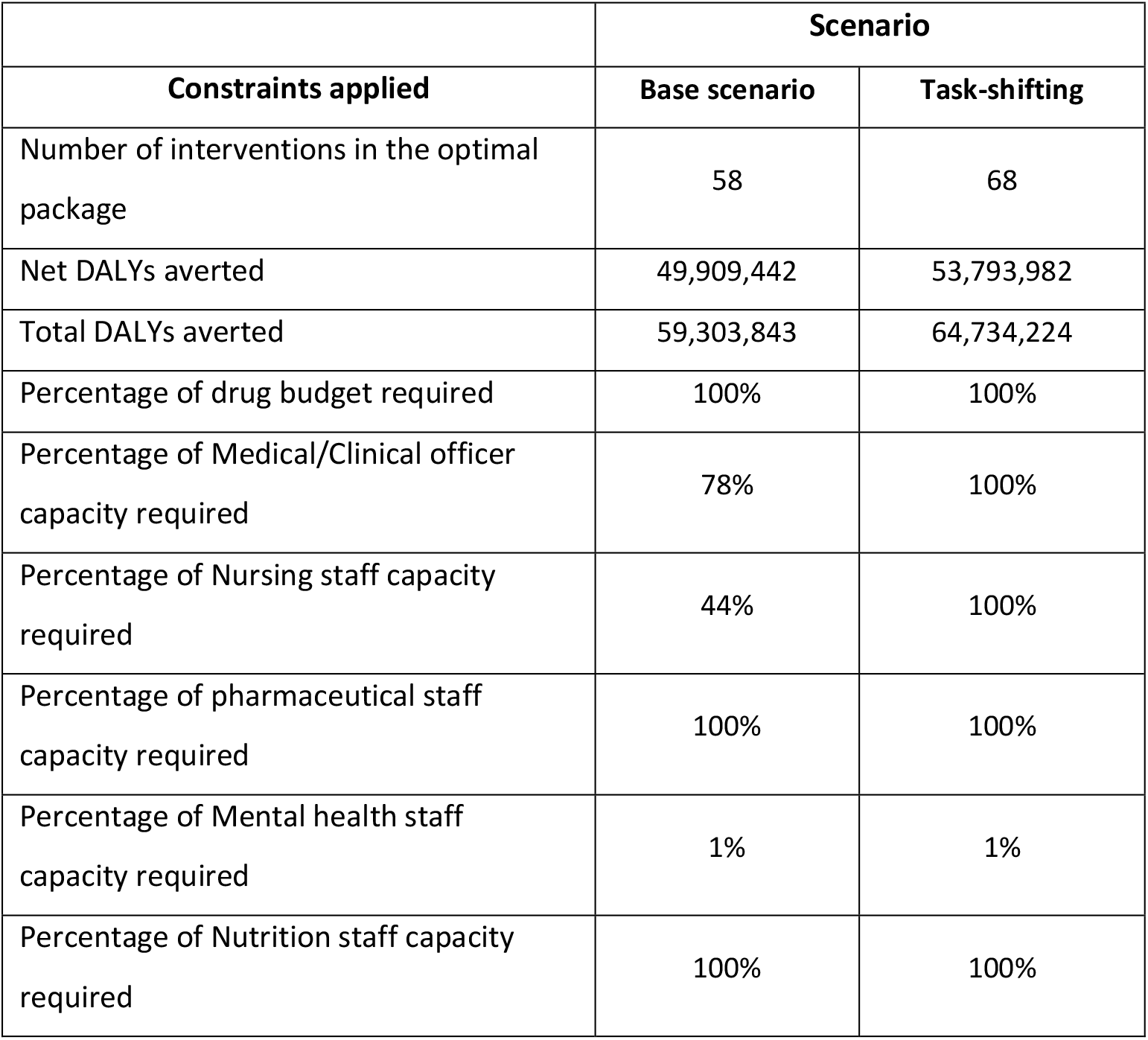
Summary of results from constrained optimization.

| Constraints applied | Scenario |  |
| --- | --- | --- |
|  | Base scenario | Task-shifting |
| Number of interventions in the optimal package | 58 | 68 |
| Net DALYs averted | 49,909,442 | 53,793,982 |
| Total DALYs averted | 59,303,843 | 64,734,224 |
| Percentage of drug budget required | 100% | 100% |
| Percentage of Medical/Clinical officer capacity required | 78% | 100% |
| Percentage of Nursing staff capacity required | 44% | 100% |
| Percentage of pharmaceutical staff capacity required | 100% | 100% |
| Percentage of Mental health staff capacity required | 1% | 1% |
| Percentage of Nutrition staff capacity required | 100% | 100% |

The above results can be further analysed by disease program, namely Reproductive, maternal, neonatal and child health (RMNCH), Non-communicable diseases (NCDs), HIV and other sexually-transmitted infections (STIs), nutrition, tuberculosis (TB), vaccine preventable diseases, malaria, mental health, integrated management of childhood illnesses (IMCI), and neglected tropical diseases (NTDs). Table 3 provides the proportion of interventions from each program included under the two scenarios. Under both scenarios, RMNCH and HIV and other STIs together account for more than half of all the interventions in the HBP. Task shifting allows more interventions to be included from five programs -RMNCH, HIV and other STIs, IMCI, nutrition and vaccine preventable diseases. Figure 1 shows the distribution of health system resource use by program and how this changes under the task-shifting scenario. By allowing task shifting, the proportional allocation of resources towards HIV and other STIs, nutrition, IMCI and vaccine preventable diseases increases, and that towards malaria, RMNCAH and TB reduces. Supplementary tables 3 and 4 provide intervention-level detail on coverage and resource use for the two scenarios.

**Table 3:** Rate of inclusion of interventions from different disease programs in the optimal HBP.

| Program | Number of interventions included in the analysis | Interventions included in the optimal package (Base scenario) |  | Interventions included in the optimal package (Task-shifting scenario) |  |
| --- | --- | --- | --- | --- | --- |
|  |  | Number | Percentage | Number | Percentage |
| RMNCH | 37 | 25 | 68% | 27 | 73% |
| NCDs | 23 | 5 | 22% | 5 | 22% |
| HIV and other STIs | 21 | 8 | 38% | 12 | 57% |
| Nutrition | 10 | 3 | 30% | 4 | 40% |
| TB | 9 | 5 | 56% | 5 | 56% |
| Vaccine Preventable Diseases | 8 | 3 | 38% | 5 | 63% |
| Malaria | 7 | 5 | 71% | 5 | 71% |
| Mental Health | 6 | 1 | 17% | 1 | 17% |
| IMCI | 4 | 1 | 25% | 2 | 50% |
| NTDs | 3 | 2 | 67% | 2 | 67% |
| **Grand Total** | **128** | **58** | **45%** | **68** | **53%** |

**Figure 1:**
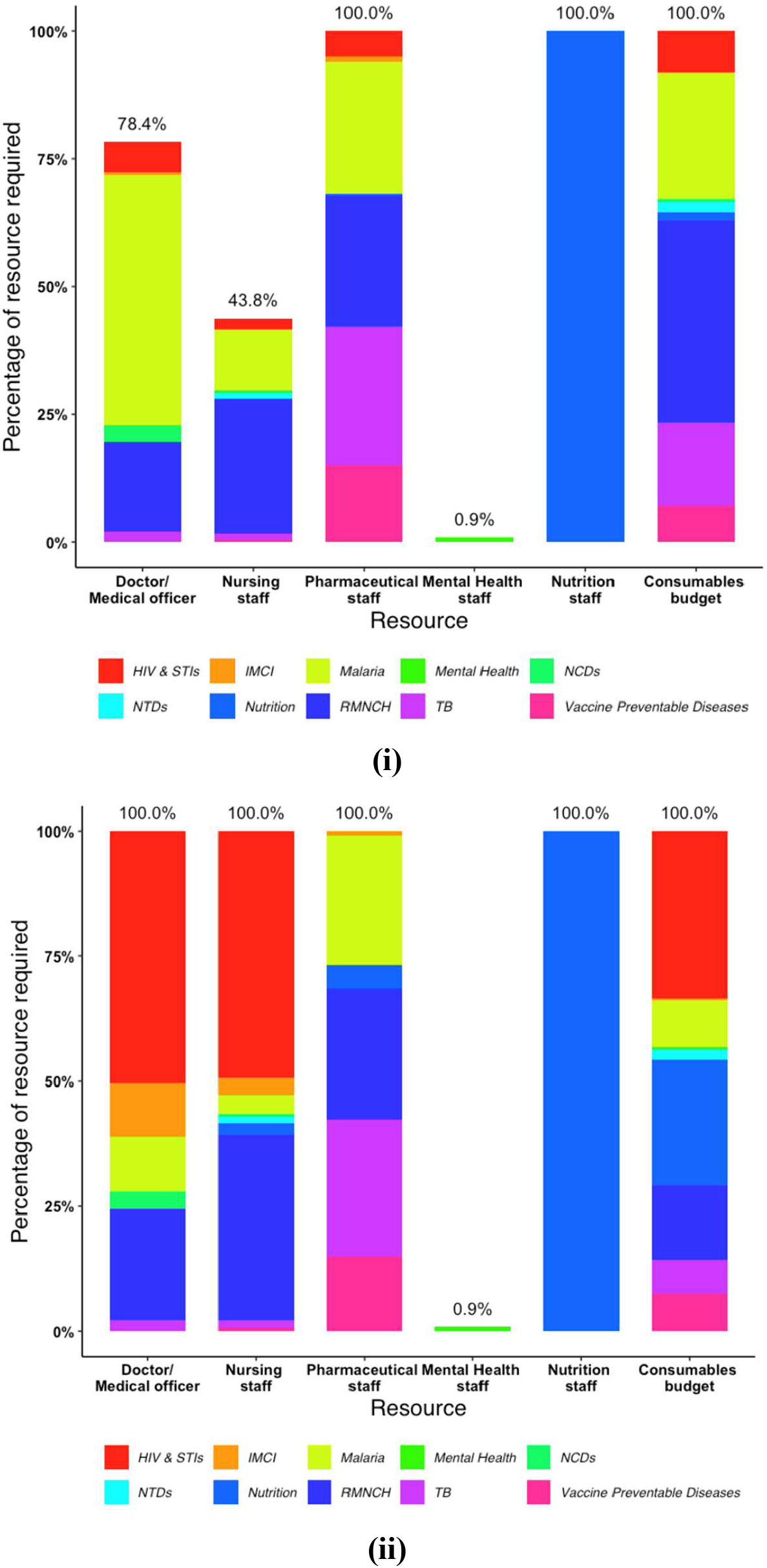
Health system resource use by program: (i) Base scenario, (ii) Task-shifting scenario: This figure illustrates the proportional distribution of health system resources across various healthcare programmes in the case of optimal resource use under the two scenarios presented in this study. (Single column fitting image)

### Effect of removing health system constraints

Supplementary table 7 demonstrates the effect of excluding the health workforce and maximum feasible coverage constraints as well as the effect of excluding the health workforce constraint alone. We note that while this omission allows for a larger HBP and health impact, the resource requirements of the resulting HBP far exceed the capacity available in Uganda and would therefore make it infeasible to implement using the current resources available.

### Marginal value of health systems components

The marginal value of relaxing health system constraints depends on the amount of additional health generated by relaxing each constraint. Figure 2 provides estimates of the net DALYs averted if an additional $1000 were spent on each of the health system components, under the two scenarios. Evidently, constraints which are not met in both scenarios will have a null marginal value (here, mental health staff). Overall, investing in health worker time provides a better outcome than expanding the consumables budget. Under both scenarios, hiring additional nutrition officer time provides the biggest impact because this helps increase the coverage of a highly cost-effective life-saving intervention - community-management of moderate acute malnutrition (MAM). Among the other health worker cadres, marginal value depends on whether task shifting to nurses is allowed. Without task shifting, pharmacists have the next highest marginal value after nutrition officers and with task shifting, the marginal value of pharmacists is superseded by that of nurses. These results can also be interpreted in terms of the cost per DALY averted. An additional $971 would need to be allocated towards the consumables budget to avert an additional DALY, net of opportunity costs. Among health workers, the cost of averting an additional net DALY is $123, $55, $56, and $39 if invested towards hiring additional time of doctors, nurses, pharmacists, and nutrition officers respectively. These estimates include the administrative costs of employing additional staff and factor in the health effects and the full costs, including downstream costs, of the interventions these health workers would be able to provide.

**Figure 2:**
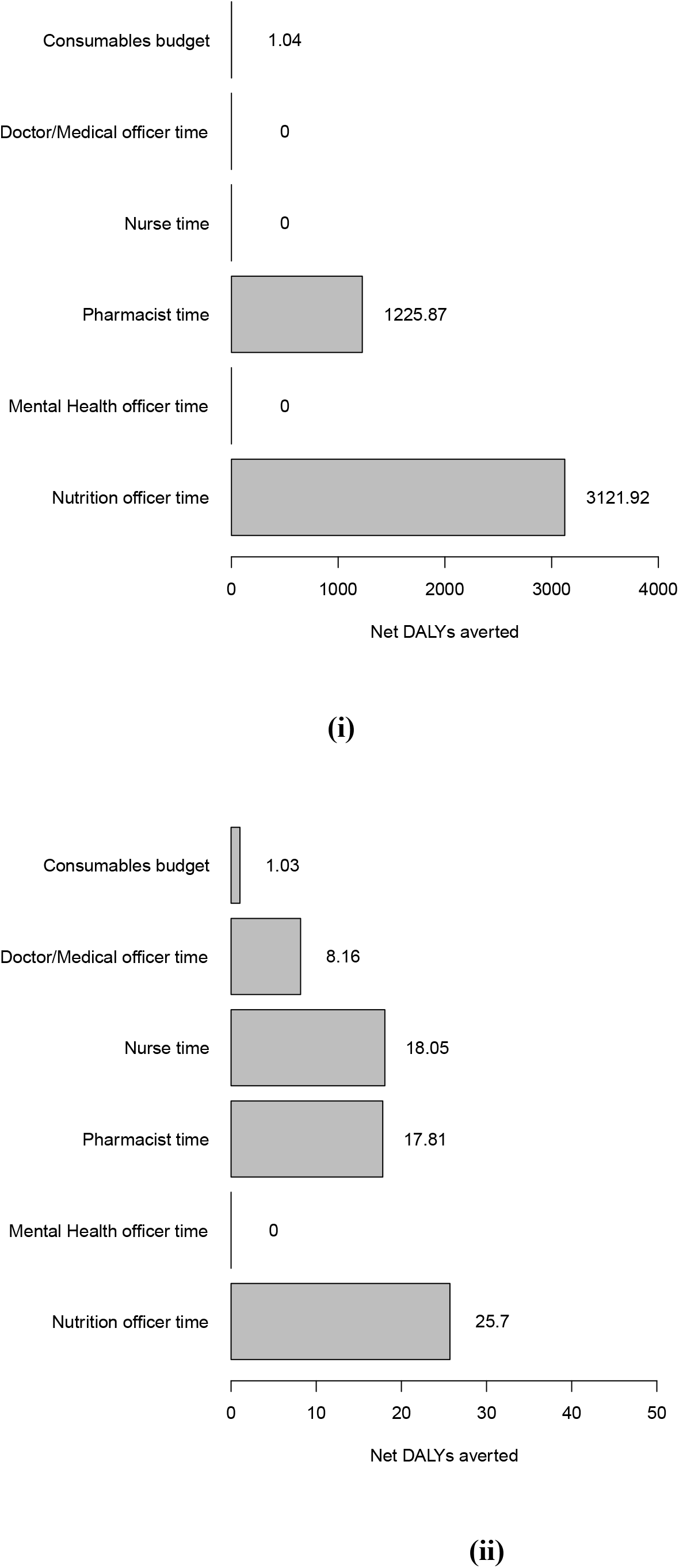
Marginal value of investing $1000 on different health system resources: (i) Base scenario (ii) Task-shifting scenario: This figure illustrates the marginal increase in net health benefit resulting from a $1000 investment towards the budget for consumables or health worker salaries by cadre under the two scenarios presented in this study. (Single column fitting image)

## Discussion

This study provides an analytical approach to inform the scope and scale of a HBP. We build upon previous work (3,5) by providing a method to explicitly account for multiple constraints which simultaneously limit the capacity of the health system to deliver services. We also show how not accounting for these constraints results in an aspirational HBP which would be infeasible to deliver. As with other approaches, the output of the above analyses is not meant to be prescriptive but rather a tool to guide decision-making as part of consultative processes(39), which demands a wider range of considerations including but not limited to the political and operational costs of adding or removing interventions from the mandate of health facilities. In the future, other objectives such as equity and financial risk protection(23) may also replace or be used in conjunction with the efficiency-focused objective of health maximization.

By providing a way to evaluate the health impact of relaxing explicitly modeled health systems constraints, our analytical framework also allows for the comparative evaluation of health system strengthening measures on the basis of their capacity to improve population health. This is an important contribution because while there have been important theoretical contributions in this area(40–42), the applied literature has been limited (26,43). Our results also demonstrate the interdependence between health systems components by showing how expanding the remit of nurses removes the bottleneck in drug dispensing and allows for a fuller use of the health systems capacity of the country. However, it is important to note that such task shifting needs to be accompanied by appropriate training and supportive supervision to avoid provision of suboptimal care.

Inevitably, this approach has some limitations. While we were able to apply the approach to a relatively data constrained setting, it is important to point out the data intensive nature of this constrained optimization methodology. The consideration of any health system resource requires information on the specific resource demands of each intervention to be evaluated in the framework. For this reason, we were able to consider only 128 interventions in our analysis, potentially excluding some efficient interventions on which evidence is limited. In the absence of adequate evidence, the inclusion of other interventions should be based on expert opinion and deliberation, followed by ‘squeezing out’ interventions from the theoretical optimal HBP to account for resources committed to these additional interventions. Related to this limitation is also the reliance on the quality of data used which can affect the quality of the final results. Another important limitation is the assumption of independence of interventions. In reality, there are likely to be complementarities and interactions between interventions as well as nonlinearity in production functions(10,16); however, quantitative evidence on this is scarce. The framework itself, however, allows for the consideration of combinations of interventions as well as incorporation of nonlinear production function through the inclusion of interventions with varying parameters at different threshold levels of coverage and may be used in this manner when better data becomes available. Furthermore, the calculation of the marginal value of investing in various health systems components assumes perfect divisibility of resources and costless transition when additional funding is added to the human resources or consumables budget. In reality, governments will need to account for administrative costs of training, hiring and procurement, and the appropriate geographic placement of additional health workers(44) among other considerations. Finally, our analysis only considers the public health sector whereas people may also be able to access care in the private sector.

Despite these limitations, we believe that our analysis serves as a useful base for the Government of Uganda to not only design their new HBP but also for other policy decisions such as health systems investments, geographic resource allocation(45), workforce training and deployment, and funding negotiations with partners. Our analysis has also demonstrated the dynamic nature of HBPs, which should change with changes in the capacity of a health system in addition to changes in epidemiology and medical technology, as well as the availability of new and better evidence, in order to allow the best use of evolving resource capabilities. In the future, we plan to apply a similar approach to assess the impact of task shifting certain primary healthcare responsibilities to relatively less trained, but more accessible community health workers.

## Data Availability

The data that supports the findings of this study are available in the supplementary material of this article.

## Ethics approval statement

The study relies upon secondary data, consisting exclusively of national level aggregate figures and does not use any personally identifiable information. No primary data collection was undertaken and secondary data were accessed and used with MOH consent.

