## Supplementary tables for "Supporting the revision of the health benefits package in Uganda: a constrained optimisation approach"

Supplementary Table 1: Data on interventions included in the constrained optimization analysis

|  |  |  | **Cost-effectiveness** | | | |  |  |  | **HR time (minutes per case)** | | | | | |
| --- | --- | --- | --- | --- | --- | --- | --- | --- | --- | --- | --- | --- | --- | --- | --- |
| **No.** | **Category** | **Intervention** | **DALYs averted per patient** | **Cost per case*** | **ICER* (cost per DALY averted)** | **Source**** | **Maximum feasible coverage (%)** | **Total number of cases in need** | **Consumables cost per case*** | **Medical Officer / Specialist** | **Nurse Officer** | **Pharmacist** | **Lab staff** | **Mental Health Staff** | **Nutrition Staff** |
| 1 | HIV & STIs | ART (Second-Line Treatment) for adults with intensive monitoring | 0.6 | $704.5 | $1093.2 | [1] | 85% | 425,461 | 1,574.3 | 4.5 | 0.0 | 0.0 | 0.0 | 0.0 | 0.0 |
| 2 | HIV & STIs | ART (Second-Line Treatment) for adults without intensive monitoring | 0.6 | $652.7 | $1,108 | [1] | 81% | 425,461 | 1,066.3 | 0.0 | 0.0 | 0.0 | 0.0 | 0.0 | 0.0 |
| 3 | HIV & STIs | ART for men | 0.6 | $211.0 | $360 | [2] | 81% | 580,675 | 152.7 | 6.0 | 4.0 | 0.5 | 0.0 | 0.0 | 0.0 |
| 4 | HIV & STIs | ART for women | 0.6 | $211.0 | $360 | [2] | 85% | 837,529 | 152.7 | 6.0 | 4.0 | 0.5 | 0.0 | 0.0 | 0.0 |
| 5 | HIV & STIs | Blood safety | 0.5 | $44.7 | $96 | [3] | 100% | 415,836 | 6.8 | 0.1 | 0.0 | 0.0 | 0.2 | 0.0 | 0.0 |
| 6 | HIV & STIs | HIV Testing Services | 0.0 | $0.8 | $45 | [1] | 75% | 21,985,282 | 14.4 | 0.0 | 0.0 | 0.0 | 0.0 | 0.0 | 0.0 |
| 7 | HIV & STIs | Home-based highly active retroviral therapy (HAART) | 6.9 | $5,541.2 | $808 | [4] | 85% | 1,418,203 | 647.6 | 4.5 | 0.0 | 0.0 | 0.0 | 0.0 | 0.0 |
| 8 | HIV & STIs | Interventions focused on male sex workers | 6.9 | $372.1 | $54 | [5] | 75% | 2,215 | 88.8 | 0.0 | 0.0 | 0.0 | 0.0 | 0.0 | 0.0 |
| 9 | HIV & STIs | Interventions focused on men who have sex with men | 6.9 | $372.1 | $54 | [5] | 75% | 24,100 | 118.4 | 0.0 | 0.0 | 0.0 | 0.0 | 0.0 | 0.0 |
| 10 | HIV & STIs | Male circumcision | 0.3 | -$94.5 | -$305 | [6] | 40% | 2,736,460 | 10.3 | 10.0 | 0.0 | 0.0 | 0.0 | 0.0 | 0.0 |
| 11 | HIV & STIs | Mass media | 0.0 | $0.0 | $2 | [1] | 71% | 41,583,600 | 0.0 | 0.0 | 0.0 | 0.0 | 0.0 | 0.0 | 0.0 |
| 12 | HIV & STIs | Peer education for sex workers | 29.5 | $68.2 | $2 | [1] | 71% | 44,300 | 0.0 | 0.0 | 0.0 | 0.0 | 0.0 | 0.0 | 0.0 |
| 13 | HIV & STIs | PMTCT | 8.6 | $334.3 | $39 | [7] | 99% | 74,517 | 22.0 | 3.0 | 10.0 | 0.5 | 0.0 | 0.0 | 0.0 |
| 14 | HIV & STIs | Pre-exposure prophylaxis for high-risk serodiscordant couples | 16.2 | $34,091.8 | $2,104 | [8] | 90% | 1,111 | 101.0 | 0.0 | 0.0 | 0.0 | 0.0 | 0.0 | 0.0 |
| 15 | HIV & STIs | Pre-exposure prophylaxis for pregnant and breastfeeding women | 0.3 | $359.3 | $1,057 | [9] | 99% | 74,517 | 101.0 | 3.0 | 10.0 | 0.5 | 0.0 | 0.0 | 0.0 |
| 16 | HIV & STIs | Screen HIV+ cases for TB | 1.0 | $114.8 | $112 | [10] | 60% | 1,676,397 | 12.0 | 4.5 | 0.0 | 0.0 | 0.0 | 0.0 | 0.0 |
| 17 | HIV & STIs | Treatment of chlamydia | 0.0 | $0.4 | $18 | [1] | 70% | 21,985,282 | 3.6 | 0.0 | 5.0 | 0.2 | 0.0 | 0.0 | 0.0 |
| 18 | HIV & STIs | Treatment of gonorrhea | 0.0 | $0.4 | $18 | [1] | 70% | 21,985,282 | 0.8 | 0.0 | 5.0 | 0.2 | 0.0 | 0.0 | 0.0 |
| 19 | HIV & STIs | Treatment of PID (Pelvic Inflammatory Disease) | 0.1 | $1.8 | $18 | [1] | 70% | 5,551,284 | 5.4 | 0.0 | 5.0 | 0.2 | 0.0 | 0.0 | 0.0 |
| 20 | HIV & STIs | Treatment of trichomoniasis | 0.0 | $0.4 | $18 | [1] | 70% | 21,985,282 | 0.4 | 0.0 | 5.0 | 0.2 | 0.0 | 0.0 | 0.0 |
| 21 | HIV & STIs | Youth focused interventions - In-school | 0.0 | $0.5 | $212 | [1] | 71% | 7,136,300 | 0.0 | 0.0 | 0.0 | 0.0 | 0.0 | 0.0 | 0.0 |
| 22 | IMCI | Antibiotics for treatment of dysentery | 0.3 | $75.0 | $250 | [11] | 46% | 905,008 | 4.9 | 5.0 | 3.5 | 0.0 | 0.0 | 0.0 | 0.0 |
| 23 | IMCI | ORS and Zinc for diarrhea | 0.0 | $3.5 | $115 | [12] | 70% | 18,272,753 | 1.1 | 5.0 | 3.5 | 0.0 | 0.0 | 0.0 | 0.0 |
| 24 | IMCI | Pneumonia treatment (children) | 0.0 | $1.0 | $48 | [13] | 71% | 10,723,306 | 0.2 | 5.0 | 3.5 | 0.0 | 0.0 | 0.0 | 0.0 |
| 25 | IMCI | Treatment of severe diarrhea | 0.6 | $58.9 | $100 | [14] | 100% | 199,339 | 2.1 | 5.0 | 3.5 | 0.0 | 0.0 | 0.0 | 0.0 |
| 26 | Malaria | Case management with artemisinin based combination therapy (80% coverage) | 0.0 | $0.3 | $21 | [15] | 100% | 25,302,308 | 2.5 | 3.5 | 1.0 | 0.0 | 0.0 | 0.0 | 0.0 |
| 27 | Malaria | Complicated (children, injectable artesunate) | 0.7 | $3.6 | $5 | [16] | 100% | 27,947 | 12.9 | 5.0 | 3.5 | 0.0 | 0.0 | 0.0 | 0.0 |
| 28 | Malaria | Home management of fevers using antimalarial (artesunate- amodiaquine AAQ) - under 5 | 3.1 | $251.4 | $81 | [17] | 87% | 2,416,001 | 2.5 | 0.0 | 3.5 | 0.0 | 0.0 | 0.0 | 0.0 |
| 29 | Malaria | Indoor residual spray and LLINs | 0.8 | $6.3 | $8 | [18] | 70% | 2,852,576 | 9.3 | 0.0 | 0.0 | 0.0 | 0.0 | 0.0 | 0.0 |
| 30 | Malaria | Intermittent preventive treatment in infants (IPTi) using 3 days of amodiaquine-artesunate (AQ3-AS3) against clinical malaria; at 2, 3, and 9 months. | 0.2 | $1.5 | $10 | [19] | 41% | 8,053,337 | 1.2 | 5.0 | 3.5 | 0.0 | 0.0 | 0.0 | 0.0 |
| 31 | Malaria | IPT (pregnant women) | 0.0 | $0.3 | $194 | [15] | 41% | 2,573,730 | 0.1 | 0.5 | 0.0 | 0.0 | 0.0 | 0.0 | 0.0 |
| 32 | Malaria | ITN distribution to pregnant women | 0.0 | $0.0 | $23 | [20] | 70% | 2,573,730 | 3.0 | 0.5 | 0.0 | 0.0 | 0.0 | 0.0 | 0.0 |
| 33 | Mental Health | Anti-epileptic medication | 0.3 | $37.1 | $121 | [21] | 10% | 196,292 | 8.0 | 0.0 | 0.0 | 0.0 | 0.0 | 15.0 | 0.0 |
| 34 | Mental Health | Basic psychosocial support, advice, and follow-up | 0.0 | $10.6 | $604 | [21] | 10% | 3,852,137 | 7.2 | 0.0 | 0.0 | 0.0 | 0.0 | 25.0 | 0.0 |
| 35 | Mental Health | Treatment of acute psychotic disorders | 0.1 | $73.7 | $1,251 | [21] | 10% | 249,502 | 18.1 | 0.0 | 0.0 | 0.0 | 0.0 | 25.0 | 0.0 |
| 36 | Mental Health | Treatment of bipolar disorder | 0.1 | $77.3 | $816 | [21] | 10% | 162,168 | 63.0 | 0.0 | 0.0 | 0.0 | 0.0 | 25.0 | 0.0 |
| 37 | Mental Health | Treatment of depression | 0.0 | $10.3 | $389 | [21] | 10% | 1,134,409 | 6.3 | 0.0 | 0.0 | 0.0 | 0.0 | 15.0 | 0.0 |
| 38 | Mental Health | Treatment of schizophrenia | 0.4 | $551.7 | $1,251 | [21] | 10% | 33,330 | 18.1 | 0.0 | 0.0 | 0.0 | 0.0 | 25.0 | 0.0 |
| 39 | NCDs | Amputation | 21.3 | $637.6 | $30 | [22] | 90% | 125 | 778.2 | 172.0 | 137.6 | 5.0 | 0.0 | 0.0 | 0.0 |
| 40 | NCDs | Asthma: Inhaled short acting beta agonist for intermittent asthma | 0.0 | $61.7 | $5,895 | [23] | 36% | 245,126 | 18.0 | 3.5 | 1.0 | 0.0 | 0.0 | 0.0 | 0.0 |
| 41 | NCDs | Asthma: Low dose inhaled beclometasone + SABA | 0.0 | $10.1 | $2,489 | [23] | 36% | 490,252 | 48.8 | 3.5 | 1.0 | 0.0 | 0.0 | 0.0 | 0.0 |
| 42 | NCDs | Breast Cancer (clinical examination + treatment) | 3.5 | $5,467.6 | $1,548 | [24] | 50% | 2,835,000 | 392.5 | 172.0 | 137.6 | 5.0 | 0.0 | 0.0 | 0.0 |
| 43 | NCDs | Breast Cancer (first line) | 0.4 | $7,418.1 | $19,239 | [24] | 10% | 13,811 | 235.7 | 172.0 | 137.6 | 5.0 | 0.0 | 0.0 | 0.0 |
| 44 | NCDs | Breast Cancer (mammography + treatment) | 0.0 | $0.5 | $993 | [25] | 50% | 1,401,400 | 397.7 | 172.0 | 137.6 | 5.0 | 0.0 | 0.0 | 0.0 |
| 45 | NCDs | Cervical cancer (first line) | 0.0 | $0.1 | $181 | [25] | 50% | 2,921 | 1,154.3 | 60.0 | 18.0 | 2.5 | 0.0 | 0.0 | 0.0 |
| 46 | NCDs | Colorectoral cancer (screening + treatment) | 0.0 | $0.3 | $266 | [25] | 10% | 213,100 | 5.3 | 172.0 | 137.6 | 5.0 | 0.0 | 0.0 | 0.0 |
| 47 | NCDs | Colorectoral cancer (treatment) | 0.0 | $0.1 | $156 | [25] | 100% | 1,111 | 680.5 | 172.0 | 137.6 | 5.0 | 0.0 | 0.0 | 0.0 |
| 48 | NCDs | community-based management of hypertension | 0.0 | $7.3 | $356 | [26] | 50% | 15,812 | 0.0 | 0.0 | 60.0 | 0.0 | 0.0 | 0.0 | 0.0 |
| 49 | NCDs | COPD - Inhaled salbutamol | 0.0 | $47.9 | $5,895 | [23] | 11% | 316,253 | 18.0 | 3.5 | 1.0 | 0.0 | 0.0 | 0.0 | 0.0 |
| 50 | NCDs | COPD - oxygen therapy and drugs | 0.0 | $200.5 | $17,186 | [23] | 5% | 37,950 | 43.2 | 5.0 | 12.0 | 2.0 | 0.0 | 0.0 | 0.0 |
| 51 | NCDs | COPD - treatment of severe exacerbations | 0.0 | $62.5 | $7,650 | [23] | 5% | 37,950 | 0.4 | 3.5 | 1.0 | 0.0 | 0.0 | 0.0 | 0.0 |
| 52 | NCDs | Elective inguinal hernia repair | 6.1 | $164.0 | $27 | [22] | 50% | 432 | 91.5 | 172.0 | 137.6 | 5.0 | 0.0 | 0.0 | 0.0 |
| 53 | NCDs | Emergency inguinal hernia repair | 35.8 | $265.9 | $7 | [22] | 90% | 728 | 91.5 | 12.0 | 12.5 | 0.0 | 0.0 | 0.0 | 0.0 |
| 54 | NCDs | GIT, Intestine cancer | 0.0 | $0.5 | $435 | [25] | 10% | 3,317,026 | 6.2 | 0.3 | 0.0 | 0.0 | 0.8 | 0.0 | 0.0 |
| 55 | NCDs | Prevention and treatment of cardiovascular disease | 0.0 | $0.3 | $54 | [27] | 5% | 3,212,775 | 30.2 | 3.5 | 1.0 | 0.0 | 0.0 | 0.0 | 0.0 |
| 56 | NCDs | Prevention of cardiovascular disease | 0.0 | $0.2 | $48 | [27] | 5% | 3,212,775 | 27.4 | 3.5 | 1.0 | 0.0 | 0.0 | 0.0 | 0.0 |
| 57 | NCDs | Retinopathy Screening and photocoagulation for diabetics | 0.0 | $1.4 | $946 | [27] | 10% | 784,741 | 1.0 | 60.0 | 18.0 | 2.5 | 0.0 | 0.0 | 0.0 |
| 58 | NCDs | Substance use disorder - alcohol | 0.0 | $18.5 | $376 | [21] | 30% | 415,836 | 0.0 | 0.0 | 60.0 | 0.0 | 0.0 | 0.0 | 0.0 |
| 59 | NCDs | Testing of pre-cancerous cells (vinegar) | 0.1 | $4.2 | $70 | [28] | 1% | 1,430,072 | 3.0 | 3.5 | 1.0 | 0.0 | 0.0 | 0.0 | 0.0 |
| 60 | NCDs | Treatment of injuries (Fracture and dislocation - fixation) | 1.2 | $452.8 | $375 | [22] | 90% | 41,584 | 24.9 | 12.0 | 12.5 | 0.0 | 0.0 | 0.0 | 0.0 |
| 61 | NCDs | Treatment of injuries (Fracture and dislocation - reduction) | 1.6 | $177.3 | $110 | [22] | 90% | 374,252 | 5.2 | 12.0 | 12.5 | 0.0 | 0.0 | 0.0 | 0.0 |
| 62 | NTDs | Schistosomiasis Mass drug administration (adults) | 0.1 | $14.8 | $122 | [29] | 80% | 6,384,460 | 0.4 | 0.0 | 0.0 | 0.0 | 0.0 | 0.0 | 0.0 |
| 63 | NTDs | Trachoma mass drug administration | 0.0 | $1.3 | $64 | [30] | 80% | 12,879,700 | 0.5 | 0.0 | 0.0 | 0.0 | 0.0 | 0.0 | 0.0 |
| 64 | NTDs | Trachoma Trichiasis cases surgey | 0.1 | $3.6 | $33 | [30] | 5% | 2,377,146 | 9.7 | 60.0 | 18.0 | 2.5 | 0.0 | 0.0 | 0.0 |
| 65 | Nutrition | Calcium Supplementation | 0.4 | $581.2 | $1,453 | [31] | 20% | 2,573,730 | 10.8 | 3.5 | 1.0 | 0.0 | 0.0 | 0.0 | 0.0 |
| 66 | Nutrition | Community management of nutrition in under-5 - micronutrient powder | 0.0 | $10.7 | $3,815 | [32] | 95% | 7,129,300 | 9.9 | 5.0 | 3.5 | 0.0 | 0.0 | 0.0 | 0.0 |
| 67 | Nutrition | Community-based management of moderate acute malnutrition (children) | 3.9 | $208.8 | $54 | [33] | 35% | 1,519,661 | 44.0 | 0.0 | 0.0 | 1.0 | 0.0 | 0.0 | 30.0 |
| 68 | Nutrition | Community-based management of severe malnutrition (children) | 3.9 | $208.8 | $54 | [33] | 35% | 1,519,661 | 130.3 | 0.0 | 0.0 | 2.0 | 0.0 | 0.0 | 40.0 |
| 69 | Nutrition | Iron fortification | 0.0 | $0.0 | $11 | [34] | 60% | 41,583,600 | 0.0 | 0.0 | 0.0 | 0.0 | 0.0 | 0.0 | 0.0 |
| 70 | Nutrition | Management of severe malnutrition (children) - inpatient | 2.4 | $609.4 | $255 | [35] | 35% | 377,065 | 94.5 | 22.5 | 137.6 | 2.0 | 0.0 | 0.0 | 210.0 |
| 71 | Nutrition | Provision of supplementary food and nutrition counselling with growth monitoring | 0.0 | $40.3 | $22,959 | [13] | 50% | 3,568,525 | 42.0 | 0.0 | 0.0 | 0.0 | 0.0 | 0.0 | 0.0 |
| 72 | Nutrition | Vitamin A supplementation in infants and children 6-59 months | 0.0 | $1.4 | $143 | [13] | 50% | 7,598,306 | 0.1 | 5.0 | 3.5 | 0.0 | 0.0 | 0.0 | 0.0 |
| 73 | Nutrition | Vitamin-A fortification (sugar) and Zinc fortification (wheat) | 0.0 | $0.1 | $11 | [13] | 95% | 7,129,300 | 0.0 | 0.0 | 0.0 | 0.0 | 0.0 | 0.0 | 0.0 |
| 74 | Nutrition | Zinc supplementation | 0.0 | $0.2 | $68 | [13] | 100% | 7,598,306 | 14.6 | 5.0 | 3.5 | 0.0 | 0.0 | 0.0 | 0.0 |
| 75 | RMNCH | Active management of the 3rd stage of labour | 7.3 | $22.6 | $3 | [11] | 55% | 1,940,089 | 0.2 | 0.5 | 0.0 | 0.0 | 0.0 | 0.0 | 0.0 |
| 76 | RMNCH | Antenatal corticosteroids for preterm labour | 1.3 | $55.4 | $44 | [36] | 20% | 97,004 | 3.4 | 0.1 | 0.0 | 0.0 | 0.0 | 0.0 | 0.0 |
| 77 | RMNCH | Antibiotics for pPRoM | 0.8 | $55.3 | $70 | [36] | 55% | 90,538 | 0.9 | 0.3 | 0.0 | 0.0 | 0.0 | 0.0 | 0.0 |
| 78 | RMNCH | Basic ANC | 0.1 | $2.1 | $26 | [37] | 100% | 2,573,730 | 38.9 | 1.5 | 30.0 | 0.0 | 0.0 | 0.0 | 0.0 |
| 79 | RMNCH | Cervical cancer screening | 0.0 | $0.3 | $352 | [25] | 70% | 4,241,284 | 0.4 | 3.5 | 1.0 | 0.0 | 0.0 | 0.0 | 0.0 |
| 80 | RMNCH | Cesearian section with indication | 9.6 | $328.6 | $34 | [22] | 92% | 13,126 | 62.9 | 22.5 | 50.0 | 5.0 | 0.0 | 0.0 | 0.0 |
| 81 | RMNCH | Cesearian Section with indication (with complication) | 27.2 | $335.3 | $12 | [22] | 92% | 2,316 | 108.3 | 22.5 | 50.0 | 5.0 | 0.0 | 0.0 | 0.0 |
| 82 | RMNCH | Chlorhexidine | 1.2 | $22.7 | $19 | [11] | 20% | 1,900,141 | 0.4 | 0.5 | 0.5 | 0.0 | 0.0 | 0.0 | 0.0 |
| 83 | RMNCH | Clean practices and immediate essential newborn care (in facility) | 0.3 | $1.4 | $5 | [38] | 76% | 1,940,089 | 1.4 | 0.1 | 0.0 | 0.0 | 0.0 | 0.0 | 0.0 |
| 84 | RMNCH | Condoms | 0.0 | $0.4 | $181 | [39] | 75% | 9,267,207 | 3.1 | 0.0 | 0.0 | 0.0 | 0.0 | 0.0 | 0.0 |
| 85 | RMNCH | Daily iron and folic acid supplementation (pregnant women) | 0.2 | $22.6 | $119 | [11] | 83% | 2,573,730 | 1.5 | 0.5 | 0.0 | 0.0 | 0.0 | 0.0 | 0.0 |
| 86 | RMNCH | Ectopic case management | 0.0 | $27.1 | $1,357 | [11] | 100% | 25,737 | 27.4 | 10.5 | 30.0 | 0.0 | 0.0 | 0.0 | 0.0 |
| 87 | RMNCH | Female sterilization | 0.4 | -$57.2 | -$155 | [40] | 73% | 351,978 | 5.0 | 60.0 | 18.0 | 2.5 | 0.0 | 0.0 | 0.0 |
| 88 | RMNCH | Fistula | 7.0 | $402.6 | $57 | [41] | 26% | 10,224 | 48.7 | 3.6 | 10.8 | 0.0 | 0.0 | 0.0 | 0.0 |
| 89 | RMNCH | Hypertensive disorder case management | 0.1 | $22.9 | $229 | [11] | 34% | 257,373 | 0.1 | 0.5 | 0.0 | 0.0 | 0.0 | 0.0 | 0.0 |
| 90 | RMNCH | Implant | 0.4 | -$57.2 | -$155 | [40] | 73% | 70,396 | 34.6 | 0.0 | 10.0 | 0.0 | 0.0 | 0.0 | 0.0 |
| 91 | RMNCH | Induction of labour (beyond 41 weeks) | 7.3 | $22.5 | $3 | [11] | 44% | 97,004 | 0.0 | 24.0 | 0.4 | 0.0 | 0.0 | 0.0 | 0.0 |
| 92 | RMNCH | Injectable | 0.4 | -$57.2 | -$155 | [40] | 73% | 3,167,798 | 5.4 | 0.0 | 10.0 | 0.0 | 0.0 | 0.0 | 0.0 |
| 93 | RMNCH | IUD | 0.4 | -$57.2 | -$155 | [40] | 73% | 281,582 | 1.7 | 0.0 | 10.0 | 0.0 | 0.0 | 0.0 | 0.0 |
| 94 | RMNCH | Kangaroo mother care | 1.8 | $22.5 | $12 | [11] | 60% | 380,028 | 0.0 | 0.0 | 0.0 | 0.0 | 0.0 | 0.0 | 0.0 |
| 95 | RMNCH | Labour and delivery management | 0.1 | $2.6 | $23 | [38] | 76% | 1,940,089 | 2.3 | 0.0 | 53.1 | 0.0 | 0.0 | 0.0 | 0.0 |
| 96 | RMNCH | Male sterilization | 0.4 | -$57.2 | -$155 | [40] | 73% | 321,920 | 8.4 | 60.0 | 18.0 | 2.5 | 0.0 | 0.0 | 0.0 |
| 97 | RMNCH | Management of eclampsia | 0.1 | $2.7 | $21 | [42] | 55% | 194,009 | 8.5 | 30.0 | 4.5 | 0.1 | 0.0 | 0.0 | 0.0 |
| 98 | RMNCH | Management of obstructed labour | 2.2 | $74.3 | $34 | [38] | 100% | 194,009 | 20.5 | 10.5 | 30.0 | 0.0 | 0.0 | 0.0 | 0.0 |
| 99 | RMNCH | Management of pre-eclampsia | 0.1 | $2.7 | $21 | [42] | 42% | 72,064 | 8.5 | 60.0 | 12.0 | 0.1 | 0.0 | 0.0 | 0.0 |
| 100 | RMNCH | Maternal Sepsis case management | 0.0 | $1.7 | $69 | [38] | 70% | 1,940,089 | 41.1 | 20.0 | 4.0 | 0.3 | 0.0 | 0.0 | 0.0 |
| 101 | RMNCH | Neonatal resuscitation (institutional) | 1.3 | $43.6 | $33 | [38] | 73% | 190,014 | 0.4 | 0.5 | 0.5 | 0.0 | 0.0 | 0.0 | 0.0 |
| 102 | RMNCH | Newborn sepsis - Full supportive care | 0.1 | $2.7 | $37 | [38] | 70% | 1,900,141 | 2.0 | 20.0 | 4.0 | 0.3 | 0.0 | 0.0 | 0.0 |
| 103 | RMNCH | Pill | 0.4 | -$57.2 | -$155 | [40] | 73% | 2,463,843 | 6.1 | 0.0 | 0.0 | 0.0 | 0.0 | 0.0 | 0.0 |
| 104 | RMNCH | Post-abortion case management | 0.1 | $28.7 | $574 | [11] | 100% | 16,950 | 15.6 | 28.0 | 28.0 | 0.3 | 0.0 | 0.0 | 0.0 |
| 105 | RMNCH | Prenatal distribution of misoprostol (for PPH prevention) | 0.1 | $0.4 | $5 | [43] | 100% | 2,573,730 | 0.4 | 0.0 | 1.0 | 0.0 | 0.0 | 0.0 | 0.0 |
| 106 | RMNCH | Safe abortion services | 0.1 | $29.1 | $485 | [11] | 3% | 339,005 | 1.3 | 15.0 | 28.0 | 0.3 | 0.0 | 0.0 | 0.0 |
| 107 | RMNCH | Support for breastfeeding mothers | 0.2 | $1.4 | $6 | [38] | 76% | 1,940,089 | 0.0 | 0.1 | 0.0 | 0.0 | 0.0 | 0.0 | 0.0 |
| 108 | RMNCH | Syphilis detection and treatment (pregnant women) | 0.1 | $0.8 | $8 | [44] | 59% | 2,573,730 | 0.5 | 0.0 | 5.0 | 0.2 | 0.0 | 0.0 | 0.0 |
| 109 | RMNCH | Tetanus toxoid (pregnant women) | 0.1 | $0.9 | $10 | [38] | 85% | 2,573,730 | 0.2 | 0.0 | 0.0 | 0.0 | 0.0 | 0.0 | 0.0 |
| 110 | RMNCH | Vaginal delivery - skilled attendance | 0.1 | $2.6 | $23 | [38] | 76% | 1,940,089 | 2.3 | 0.0 | 53.1 | 0.0 | 0.0 | 0.0 | 0.0 |
| 111 | RMNCH | Vaginal Delivery - with complication | 0.6 | $12.9 | $23 | [38] | 92% | 383,743 | 18.6 | 10.5 | 30.0 | 0.0 | 0.0 | 0.0 | 0.0 |
| 112 | TB | Cotrimoxazole preventive therapy for TB HIV+ patients | 0.1 | $16.1 | $234 | [45] | 41% | 8,381,990 | 9.3 | 2.5 | 11.3 | 1.5 | 0.0 | 0.0 | 0.0 |
| 113 | TB | First line treatment of smear positive cases (95% coverage) | 72.3 | $325.5 | $5 | [46] | 85% | 46,043 | 37.3 | 4.5 | 8.0 | 0.0 | 0.0 | 0.0 | 0.0 |
| 114 | TB | Full combination DOTS (smear-positive, smear negative, extrapulmonary cases, MDR cases) | 78.7 | $662.1 | $8 | [46] | 33% | 48,201 | 4,376.0 | 2.5 | 11.3 | 1.5 | 0.0 | 0.0 | 0.0 |
| 115 | TB | Full DOTS (smear-positive, smear negative and Extrapulmonary cases) | 78.7 | $560.2 | $7 | [46] | 33% | 47,170 | 622.1 | 2.5 | 11.3 | 1.5 | 0.0 | 0.0 | 0.0 |
| 116 | TB | Isonized Preventive Therapy for children in contact with TB patients | 0.5 | $48.9 | $104 | [47] | 85% | 46,037 | 2.2 | 4.5 | 8.0 | 0.0 | 0.0 | 0.0 | 0.0 |
| 117 | TB | Isonized Preventive Therapy for HIV+ people | 10.8 | $12.0 | $1 | [48] | 85% | 1,380,000 | 11.3 | 4.5 | 8.0 | 0.0 | 0.0 | 0.0 | 0.0 |
| 118 | TB | Isonized Preventive Therapy for HIV+ pregnant women | 20.3 | $24.3 | $1 | [49] | 85% | 127,330 | 11.3 | 4.5 | 8.0 | 0.0 | 0.0 | 0.0 | 0.0 |
| 119 | TB | Xpert test (Full) | 0.0 | $0.6 | $137 | [50] | 90% | 781,221 | 11.5 | 0.0 | 0.0 | 0.0 | 0.0 | 0.0 | 0.0 |
| 120 | TB | Xpert test (targeted) | 0.0 | $1.1 | $217 | [50] | 90% | 377,881 | 11.5 | 0.0 | 0.0 | 0.0 | 0.0 | 0.0 | 0.0 |
| 121 | Vaccine Preventable Diseases | BCG vaccine | 0.1 | $13.1 | $114 | [51] | 90% | 1,861,758 | 0.6 | 0.0 | 0.0 | 0.0 | 0.0 | 0.0 | 0.0 |
| 122 | Vaccine Preventable Diseases | DPT-Heb-Hib / Pentavalent vaccine | 4.2 | $212.0 | $51 | [52] | 90% | 1,861,758 | 1.8 | 0.0 | 0.0 | 0.0 | 0.0 | 0.0 | 0.0 |
| 123 | Vaccine Preventable Diseases | HPV vaccine | 0.0 | $6.4 | $196 | [28] | 80% | 649,908 | 9.1 | 0.0 | 0.0 | 0.0 | 0.0 | 0.0 | 0.0 |
| 124 | Vaccine Preventable Diseases | Measles vaccine | 0.0 | $0.0 | $3 | [53] | 90% | 1,861,758 | 0.7 | 0.0 | 0.0 | 0.0 | 0.0 | 0.0 | 0.0 |
| 125 | Vaccine Preventable Diseases | Oral cholera vaccine | 0.0 | $2.5 | $1,333 | [54] | 53% | 4,854,823 | 2.7 | 0.0 | 0.0 | 0.0 | 0.0 | 0.0 | 0.0 |
| 126 | Vaccine Preventable Diseases | Pneumococcal vaccine | 3.5 | $261.7 | $75 | [55] | 90% | 1,861,758 | 9.3 | 0.0 | 0.0 | 0.0 | 0.0 | 0.0 | 0.0 |
| 127 | Vaccine Preventable Diseases | Polio vaccine | 0.0 | $1.0 | $2,040 | [56] | 90% | 1,861,758 | 0.5 | 0.0 | 0.0 | 0.0 | 0.0 | 0.0 | 0.0 |
| 128 | Vaccine Preventable Diseases | Rotavirus vaccine | 0.2 | $11.4 | $61 | [57] | 90% | 1,861,758 | 4.6 | 0.0 | 0.0 | 0.0 | 0.0 | 0.0 | 0.0 |

*All monetary figures in 2019 US$

** References for cost-effectiveness evidence:

| [1]: | Hogan et al. (2005) |
| --- | --- |
| [2]: | Alistar et al. (2014) |
| [3]: | Aldridge (2009) |
| [4]: | Marseille (2009) |
| [5]: | Vassall (2014) |
| [6]: | Uthman (2011) |
| [7]: | Kuznik (2012) |
| [8]: | Ying (2015) |
| [9]: | Price (2016) |
| [10]: | Shah (2014) |
| [11]: | WHO-CHOICE |
| [12]: | Robberstad (2004) |
| [13]: | Edejer et al. (2005) |
| [14]: | Ruhago (2015) |
| [15]: | Morel (2005) |
| [16]: | Lubell (2011) |
| [17]: | Nonvignon (2012) |
| [18]: | Stuckey (2014) |
| [19]: | Conteh (2010) |
| [20]: | Becker-Dreps (2009) |
| [21]: | Chisholm (2012) |
| [22]: | Roberts (2016) |
| [23]: | Stanciole (2012) |
| [24]: | Zelle (2012) |
| [25]: | Ginsberg (2012) |
| [26]: | Gaziano (2014) |
| [27]: | Ortegón (2012) |
| [28]: | Campos (2017) |
| [29]: | Lo (2015) |
| [30]: | Baltussen (2012) |
| [31]: | Robberstad (2007) |
| [32]: | Pasricha et al. (2020) |
| [33]: | Wilford (2012) |
| [34]: | Baltussen et al. (2004) |
| [35]: | Puett (2013) |
| [36]: | Darmstadt et al, 2005 |
| [37]: | Darmstadt, 2004 |
| [38]: | Adam (2005) |
| [39]: | Stover (2017) |
| [40]: | Babigumira (2012) |
| [41]: | Epiu (2018) |
| [42]: | Feldhaus (2016) |
| [43]: | Lubinga et al. (2016) |
| [44]: | Kuznik (2013) |
| [45]: | Pitter (2007) |
| [46]: | Baltussen (2005) |
| [47]: | Jo et al. (2021) |
| [48]: | Johnson (2018) |
| [49]: | Kapoor (2016) |
| [50]: | Tesfaye (2017) |
| [51]: | Machlaurin (2020) |
| [52]: | Akumu (2007) |
| [53]: | Kaucley (2015) |
| [54]: | Jeuland (2009) |
| [55]: | Sinha (2007) |
| [56]: | Khan (2008) |
| [57]: | Kim (2011) |

Supplementary Table 2: Optimum coverage level of various interventions under the base scenario and task-shifting scenario

| **No.** | **Program** | **Intervention** | **Base scenario** | **Task-shifting scenario** |
| --- | --- | --- | --- | --- |
| 1 | HIV & STIs | ART (Second-Line Treatment) for adults with intensive monitoring | 0% | 0% |
| 2 | HIV & STIs | ART (Second-Line Treatment) for adults without intensive monitoring | 0% | 0% |
| 3 | HIV & STIs | ART for men | 0% | 0% |
| 4 | HIV & STIs | ART for women | 0% | 0% |
| 5 | HIV & STIs | Blood safety | 100% | 100% |
| 6 | HIV & STIs | HIV Testing Services | 0% | 0% |
| 7 | HIV & STIs | Home-based highly active retroviral therapy (HAART) | 0% | 0% |
| 8 | HIV & STIs | Interventions focused on male sex workers | 75% | 75% |
| 9 | HIV & STIs | Interventions focused on men who have sex with men | 75% | 75% |
| 10 | HIV & STIs | Male circumcision | 40% | 40% |
| 11 | HIV & STIs | Mass media | 71% | 71% |
| 12 | HIV & STIs | Peer education for sex workers | 71% | 71% |
| 13 | HIV & STIs | PMTCT | 99% | 99% |
| 14 | HIV & STIs | Pre-exposure prophylaxis for high-risk serodiscordant couples | 0% | 0% |
| 15 | HIV & STIs | Pre-exposure prophylaxis for pregnant and breastfeeding women | 0% | 0% |
| 16 | HIV & STIs | Screen HIV+ cases for TB | 60% | 60% |
| 17 | HIV & STIs | Treatment of chlamydia | 0% | 70% |
| 18 | HIV & STIs | Treatment of gonorrhea | 0% | 70% |
| 19 | HIV & STIs | Treatment of PID (Pelvic Inflammatory Disease) | 0% | 70% |
| 20 | HIV & STIs | Treatment of trichomoniasis | 0% | 70% |
| 21 | HIV & STIs | Youth focused interventions - In-school | 0% | 0% |
| 22 | IMCI | Antibiotics for treatment of dysentery | 0% | 0% |
| 23 | IMCI | ORS and Zinc for diarrhea | 0% | 0% |
| 24 | IMCI | Pneumonia treatment (children) | 0% | 49% |
| 25 | IMCI | Treatment of severe diarrhea | 100% | 100% |
| 26 | Malaria | Case management with artemisinin based combination therapy (80% coverage) | 100% | 11% |
| 27 | Malaria | Complicated (children, injectable artesunate) | 100% | 100% |
| 28 | Malaria | Home management of fevers using antimalarial (artesunate- amodiaquine AAQ) - under 5 | 87% | 87% |
| 29 | Malaria | Indoor residual spray and LLINs | 70% | 70% |
| 30 | Malaria | Intermittent preventive treatment in infants (IPTi) using 3 days of amodiaquine-artesunate (AQ3-AS3) against clinical malaria; at 2, 3, and 9 months. | 41% | 41% |
| 31 | Malaria | IPT (pregnant women) | 0% | 0% |
| 32 | Malaria | ITN distribution to pregnant women | 0% | 0% |
| 33 | Mental Health | Anti-epileptic medication | 10% | 10% |
| 34 | Mental Health | Basic psychosocial support, advice, and follow-up | 0% | 0% |
| 35 | Mental Health | Treatment of acute psychotic disorders | 0% | 0% |
| 36 | Mental Health | Treatment of bipolar disorder | 0% | 0% |
| 37 | Mental Health | Treatment of depression | 0% | 0% |
| 38 | Mental Health | Treatment of schizophrenia | 0% | 0% |
| 39 | NCDs | Amputation | 90% | 90% |
| 40 | NCDs | Asthma: Inhaled short acting beta agonist for intermittent asthma | 0% | 0% |
| 41 | NCDs | Asthma: Low dose inhaled beclometasone + SABA | 0% | 0% |
| 42 | NCDs | Breast Cancer (clinical examination + treatment) | 0% | 0% |
| 43 | NCDs | Breast Cancer (first line) | 0% | 0% |
| 44 | NCDs | Breast Cancer (mammography + treatment) | 0% | 0% |
| 45 | NCDs | Cervical cancer (first line) | 0% | 0% |
| 46 | NCDs | Colorectoral cancer (screening + treatment) | 0% | 0% |
| 47 | NCDs | Colorectoral cancer (treatment) | 0% | 0% |
| 48 | NCDs | community-based management of hypertension | 0% | 0% |
| 49 | NCDs | COPD - Inhaled salbutamol | 0% | 0% |
| 50 | NCDs | COPD - oxygen therapy and drugs | 0% | 0% |
| 51 | NCDs | COPD - treatment of severe exacerbations | 0% | 0% |
| 52 | NCDs | Elective inguinal hernia repair | 50% | 50% |
| 53 | NCDs | Emergency inguinal hernia repair | 90% | 90% |
| 54 | NCDs | GIT, Intestine cancer | 0% | 0% |
| 55 | NCDs | Prevention and treatment of cardiovascular disease | 0% | 0% |
| 56 | NCDs | Prevention of cardiovascular disease | 0% | 0% |
| 57 | NCDs | Retinopathy Screening and photocoagulation for diabetics | 0% | 0% |
| 58 | NCDs | Substance use disorder - alcohol | 0% | 0% |
| 59 | NCDs | Testing of pre-cancerous cells (vinegar) | 1% | 1% |
| 60 | NCDs | Treatment of injuries (Fracture and dislocation - fixation) | 0% | 0% |
| 61 | NCDs | Treatment of injuries (Fracture and dislocation - reduction) | 90% | 90% |
| 62 | NTDs | Schistosomiasis Mass drug administration (adults) | 80% | 80% |
| 63 | NTDs | Trachoma mass drug administration | 80% | 80% |
| 64 | NTDs | Trachoma Trichiasis cases surgey | 0% | 0% |
| 65 | Nutrition | Calcium Supplementation | 0% | 0% |
| 66 | Nutrition | Community management of nutrition in under-5 - micronutrient powder | 0% | 0% |
| 67 | Nutrition | Community-based management of moderate acute malnutrition (children) | 7% | 35% |
| 68 | Nutrition | Community-based management of severe malnutrition (children) | 0% | 35% |
| 69 | Nutrition | Iron fortification | 60% | 60% |
| 70 | Nutrition | Management of severe malnutrition (children) - inpatient | 0% | 0% |
| 71 | Nutrition | Provision of supplementary food and nutrition counselling with growth monitoring | 0% | 0% |
| 72 | Nutrition | Vitamin A supplementation in infants and children 6-59 months | 0% | 0% |
| 73 | Nutrition | Vitamin-A fortification (sugar) and Zinc fortification (wheat) | 95% | 95% |
| 74 | Nutrition | Zinc supplementation | 0% | 0% |
| 75 | RMNCH | Active management of the 3rd stage of labour | 55% | 55% |
| 76 | RMNCH | Antenatal corticosteroids for preterm labour | 20% | 20% |
| 77 | RMNCH | Antibiotics for pPRoM | 55% | 55% |
| 78 | RMNCH | Basic ANC | 100% | 0% |
| 79 | RMNCH | Cervical cancer screening | 0% | 0% |
| 80 | RMNCH | Cesearian section with indication | 92% | 92% |
| 81 | RMNCH | Cesearian Section with indication (with complication) | 92% | 92% |
| 82 | RMNCH | Chlorhexidine | 20% | 20% |
| 83 | RMNCH | Clean practices and immediate essential newborn care (in facility) | 76% | 76% |
| 84 | RMNCH | Condoms | 0% | 0% |
| 85 | RMNCH | Daily iron and folic acid supplementation (pregnant women) | 83% | 83% |
| 86 | RMNCH | Ectopic case management | 0% | 0% |
| 87 | RMNCH | Female sterilization | 73% | 73% |
| 88 | RMNCH | Fistula | 26% | 26% |
| 89 | RMNCH | Hypertensive disorder case management | 0% | 0% |
| 90 | RMNCH | Implant | 73% | 73% |
| 91 | RMNCH | Induction of labour (beyond 41 weeks) | 44% | 44% |
| 92 | RMNCH | Injectable | 73% | 73% |
| 93 | RMNCH | IUD | 73% | 73% |
| 94 | RMNCH | Kangaroo mother care | 60% | 60% |
| 95 | RMNCH | Labour and delivery management | 10% | 26% |
| 96 | RMNCH | Male sterilization | 73% | 73% |
| 97 | RMNCH | Management of eclampsia | 0% | 0% |
| 98 | RMNCH | Management of obstructed labour | 100% | 100% |
| 99 | RMNCH | Management of pre-eclampsia | 0% | 0% |
| 100 | RMNCH | Maternal Sepsis case management | 0% | 0% |
| 101 | RMNCH | Neonatal resuscitation (institutional) | 73% | 73% |
| 102 | RMNCH | Newborn sepsis - Full supportive care | 0% | 70% |
| 103 | RMNCH | Pill | 73% | 73% |
| 104 | RMNCH | Post-abortion case management | 0% | 0% |
| 105 | RMNCH | Prenatal distribution of misoprostol (for PPH prevention) | 100% | 100% |
| 106 | RMNCH | Safe abortion services | 0% | 0% |
| 107 | RMNCH | Support for breastfeeding mothers | 76% | 76% |
| 108 | RMNCH | Syphilis detection and treatment (pregnant women) | 0% | 59% |
| 109 | RMNCH | Tetanus toxoid (pregnant women) | 85% | 85% |
| 110 | RMNCH | Vaginal delivery - skilled attendance | 0% | 76% |
| 111 | RMNCH | Vaginal Delivery - with complication | 92% | 92% |
| 112 | TB | Cotrimoxazole preventive therapy for TB HIV+ patients | 0% | 0% |
| 113 | TB | First line treatment of smear positive cases (95% coverage) | 33% | 51% |
| 114 | TB | Full combination DOTS (smear-positive, smear negative, extrapulmonary cases, MDR cases) | 17% | 0% |
| 115 | TB | Full DOTS (smear-positive, smear negative and Extrapulmonary cases) | 33% | 33% |
| 116 | TB | Isonized Preventive Therapy for children in contact with TB patients | 0% | 85% |
| 117 | TB | Isonized Preventive Therapy for HIV+ people | 85% | 85% |
| 118 | TB | Isonized Preventive Therapy for HIV+ pregnant women | 85% | 85% |
| 119 | TB | Xpert test (Full) | 0% | 0% |
| 120 | TB | Xpert test (targeted) | 0% | 0% |
| 121 | Vaccine Preventable Diseases | BCG vaccine | 0% | 90% |
| 122 | Vaccine Preventable Diseases | DPT-Heb-Hib / Pentavalent vaccine | 90% | 90% |
| 123 | Vaccine Preventable Diseases | HPV vaccine | 0% | 0% |
| 124 | Vaccine Preventable Diseases | Measles vaccine | 0% | 90% |
| 125 | Vaccine Preventable Diseases | Oral cholera vaccine | 0% | 0% |
| 126 | Vaccine Preventable Diseases | Pneumococcal vaccine | 90% | 90% |
| 127 | Vaccine Preventable Diseases | Polio vaccine | 0% | 0% |
| 128 | Vaccine Preventable Diseases | Rotavirus vaccine | 90% | 90% |

Supplementary Table 3: Health outcomes and resource use by intervention under the base scenario

| **No.** | **Program** | **Intervention** | **% of cases in need covered under optimal package** | **Total number of cases covered under the optimal package** | **DALYs averted** | **Consumables expenditure required** | **Medical/Clinical Officer** | **Nursing staff** | **Pharmaceutical staff** | **Lab staff** | **Mental health staff** | **Nutrition staff** |
| --- | --- | --- | --- | --- | --- | --- | --- | --- | --- | --- | --- | --- |
| 1 | HIV & STIs | ART (Second-Line Treatment) for adults with intensive monitoring | 0% | - | - | - | 0 | 0 | 0 | 0 | 0 | 0 |
| 2 | HIV & STIs | ART (Second-Line Treatment) for adults without intensive monitoring | 0% | - | - | - | 0 | 0 | 0 | 0 | 0 | 0 |
| 3 | HIV & STIs | ART for men | 0% | - | - | - | 0 | 0 | 0 | 0 | 0 | 0 |
| 4 | HIV & STIs | ART for women | 0% | - | - | - | 0 | 0 | 0 | 0 | 0 | 0 |
| 5 | HIV & STIs | Blood safety | 100% | 415,836 | 193,120 | $2,813,131 | 1 | 0 | 0 | 10 | 0 | 0 |
| 6 | HIV & STIs | HIV Testing Services | 0% | - | - | - | 0 | 0 | 0 | 0 | 0 | 0 |
| 7 | HIV & STIs | Home-based highly active retroviral therapy (HAART) | 0% | - | - | - | 0 | 0 | 0 | 0 | 0 | 0 |
| 8 | HIV & STIs | Interventions focused on male sex workers | 75% | 1,661 | 11,413 | $147,500 | 0 | 1 | 0 | 0 | 0 | 0 |
| 9 | HIV & STIs | Interventions focused on men who have sex with men | 75% | 18,075 | 124,175 | $2,139,638 | 0 | 12 | 0 | 0 | 0 | 0 |
| 10 | HIV & STIs | Male circumcision | 40% | 1,094,584 | 339,321 | $11,283,038 | 313 | 420 | 0 | 0 | 0 | 0 |
| 11 | HIV & STIs | Mass media | 71% | 29,524,356 | 250,311 | $0 | 0 | 0 | 0 | 0 | 0 | 0 |
| 12 | HIV & STIs | Peer education for sex workers | 71% | 31,453 | 928,931 | $0 | 0 | 0 | 0 | 0 | 0 | 0 |
| 13 | HIV & STIs | PMTCT | 99% | 73,586 | 631,366 | $1,615,645 | 6 | 28 | 3 | 0 | 0 | 0 |
| 14 | HIV & STIs | Pre-exposure prophylaxis for high-risk serodiscordant couples | 0% | - | - | - | 0 | 0 | 0 | 0 | 0 | 0 |
| 15 | HIV & STIs | Pre-exposure prophylaxis for pregnant and breastfeeding women | 0% | - | - | - | 0 | 0 | 0 | 0 | 0 | 0 |
| 16 | HIV & STIs | Screen HIV+ cases for TB | 60% | 1,005,838 | 1,028,700 | $12,037,082 | 173 | 116 | 21 | 0 | 0 | 0 |
| 17 | HIV & STIs | Treatment of chlamydia | 0% | - | - | - | 0 | 0 | 0 | 0 | 0 | 0 |
| 18 | HIV & STIs | Treatment of gonorrhea | 0% | - | - | - | 0 | 0 | 0 | 0 | 0 | 0 |
| 19 | HIV & STIs | Treatment of PID (Pelvic Inflammatory Disease) | 0% | - | - | - | 0 | 0 | 0 | 0 | 0 | 0 |
| 20 | HIV & STIs | Treatment of trichomoniasis | 0% | - | - | - | 0 | 0 | 0 | 0 | 0 | 0 |
| 21 | HIV & STIs | Youth focused interventions - In-school | 0% | - | - | - | 0 | 0 | 0 | 0 | 0 | 0 |
| 22 | IMCI | Antibiotics for treatment of dysentery | 0% | - | - | - | 0 | 0 | 0 | 0 | 0 | 0 |
| 23 | IMCI | ORS and Zinc for diarrhea | 0% | - | - | - | 0 | 0 | 0 | 0 | 0 | 0 |
| 24 | IMCI | Pneumonia treatment (children) | 0% | - | - | - | 0 | 0 | 0 | 0 | 0 | 0 |
| 25 | IMCI | Treatment of severe diarrhea | 100% | 199,339 | 117,610 | $416,064 | 31 | 27 | 4 | 0 | 0 | 0 |
| 26 | Malaria | Case management with artemisinin based combination therapy (80% coverage) | 100% | 25,302,308 | 396,066 | $64,344,608 | 3438 | 2425 | 0 | 0 | 0 | 0 |
| 27 | Malaria | Complicated (children, injectable artesunate) | 100% | 27,947 | 19,563 | $360,158 | 4 | 4 | 1 | 0 | 0 | 0 |
| 28 | Malaria | Home management of fevers using antimalarial (artesunate- amodiaquine AAQ) - under 5 | 87% | 2,101,921 | 6,499,956 | $5,345,255 | 0 | 282 | 47 | 0 | 0 | 0 |
| 29 | Malaria | Indoor residual spray and LLINs | 70% | 1,996,803 | 1,522,962 | $18,470,427 | 0 | 38 | 0 | 0 | 0 | 0 |
| 30 | Malaria | Intermittent preventive treatment in infants (IPTi) using 3 days of amodiaquine-artesunate (AQ3-AS3) against clinical malaria; at 2, 3, and 9 months. | 41% | 3,301,868 | 521,695 | $4,107,524 | 519 | 443 | 74 | 0 | 0 | 0 |
| 31 | Malaria | IPT (pregnant women) | 0% | - | - | - | 0 | 0 | 0 | 0 | 0 | 0 |
| 32 | Malaria | ITN distribution to pregnant women | 0% | - | - | - | 0 | 0 | 0 | 0 | 0 | 0 |
| 33 | Mental Health | Anti-epileptic medication | 10% | 19,629 | 6,021 | $156,164 | 0 | 0 | 0 | 0 | 4 | 0 |
| 34 | Mental Health | Basic psychosocial support, advice, and follow-up | 0% | - | - | - | 0 | 0 | 0 | 0 | 0 | 0 |
| 35 | Mental Health | Treatment of acute psychotic disorders | 0% | - | - | - | 0 | 0 | 0 | 0 | 0 | 0 |
| 36 | Mental Health | Treatment of bipolar disorder | 0% | - | - | - | 0 | 0 | 0 | 0 | 0 | 0 |
| 37 | Mental Health | Treatment of depression | 0% | - | - | - | 0 | 0 | 0 | 0 | 0 | 0 |
| 38 | Mental Health | Treatment of schizophrenia | 0% | - | - | - | 0 | 0 | 0 | 0 | 0 | 0 |
| 39 | NCDs | Amputation | 90% | 112 | 2,387 | $87,375 | 1 | 0 | 0 | 0 | 0 | 0 |
| 40 | NCDs | Asthma: Inhaled short acting beta agonist for intermittent asthma | 0% | - | - | - | 0 | 0 | 0 | 0 | 0 | 0 |
| 41 | NCDs | Asthma: Low dose inhaled beclometasone + SABA | 0% | - | - | - | 0 | 0 | 0 | 0 | 0 | 0 |
| 42 | NCDs | Breast Cancer (clinical examination + treatment) | 0% | - | - | - | 0 | 0 | 0 | 0 | 0 | 0 |
| 43 | NCDs | Breast Cancer (first line) | 0% | - | - | - | 0 | 0 | 0 | 0 | 0 | 0 |
| 44 | NCDs | Breast Cancer (mammography + treatment) | 0% | - | - | - | 0 | 0 | 0 | 0 | 0 | 0 |
| 45 | NCDs | Cervical cancer (first line) | 0% | - | - | - | 0 | 0 | 0 | 0 | 0 | 0 |
| 46 | NCDs | Colorectoral cancer (screening + treatment) | 0% | - | - | - | 0 | 0 | 0 | 0 | 0 | 0 |
| 47 | NCDs | Colorectoral cancer (treatment) | 0% | - | - | - | 0 | 0 | 0 | 0 | 0 | 0 |
| 48 | NCDs | community-based management of hypertension | 0% | - | - | - | 0 | 0 | 0 | 0 | 0 | 0 |
| 49 | NCDs | COPD - Inhaled salbutamol | 0% | - | - | - | 0 | 0 | 0 | 0 | 0 | 0 |
| 50 | NCDs | COPD - oxygen therapy and drugs | 0% | - | - | - | 0 | 0 | 0 | 0 | 0 | 0 |
| 51 | NCDs | COPD - treatment of severe exacerbations | 0% | - | - | - | 0 | 0 | 0 | 0 | 0 | 0 |
| 52 | NCDs | Elective inguinal hernia repair | 50% | 216 | 1,322 | $19,754 | 1 | 1 | 0 | 0 | 0 | 0 |
| 53 | NCDs | Emergency inguinal hernia repair | 90% | 655 | 23,469 | $59,947 | 1 | 0 | 0 | 0 | 0 | 0 |
| 54 | NCDs | GIT, Intestine cancer | 0% | - | - | - | 0 | 0 | 0 | 0 | 0 | 0 |
| 55 | NCDs | Prevention and treatment of cardiovascular disease | 0% | - | - | - | 0 | 0 | 0 | 0 | 0 | 0 |
| 56 | NCDs | Prevention of cardiovascular disease | 0% | - | - | - | 0 | 0 | 0 | 0 | 0 | 0 |
| 57 | NCDs | Retinopathy Screening and photocoagulation for diabetics | 0% | - | - | - | 0 | 0 | 0 | 0 | 0 | 0 |
| 58 | NCDs | Substance use disorder - alcohol | 0% | - | - | - | 0 | 0 | 0 | 0 | 0 | 0 |
| 59 | NCDs | Testing of pre-cancerous cells (vinegar) | 1% | 11,172 | 674 | $33,407 | 2 | 1 | 0 | 0 | 0 | 0 |
| 60 | NCDs | Treatment of injuries (Fracture and dislocation - fixation) | 0% | - | - | - | 0 | 0 | 0 | 0 | 0 | 0 |
| 61 | NCDs | Treatment of injuries (Fracture and dislocation - reduction) | 90% | 374,252 | 603,164 | $1,946,112 | 300 | 179 | 0 | 0 | 0 | 0 |
| 62 | NTDs | Schistosomiasis Mass drug administration (adults) | 80% | 5,107,568 | 620,391 | $1,833,701 | 0 | 98 | 0 | 0 | 0 | 0 |
| 63 | NTDs | Trachoma mass drug administration | 80% | 10,303,760 | 208,421 | $5,467,939 | 0 | 197 | 0 | 0 | 0 | 0 |
| 64 | NTDs | Trachoma Trichiasis cases surgey | 0% | - | - | - | 0 | 0 | 0 | 0 | 0 | 0 |
| 65 | Nutrition | Calcium Supplementation | 0% | - | - | - | 0 | 0 | 0 | 0 | 0 | 0 |
| 66 | Nutrition | Community management of nutrition in under-5 - micronutrient powder | 0% | - | - | - | 0 | 0 | 0 | 0 | 0 | 0 |
| 67 | Nutrition | Community-based management of moderate acute malnutrition (children) | 7% | 113,963 | 444,456 | $5,010,851 | 0 | 0 | 2 | 0 | 0 | 46 |
| 68 | Nutrition | Community-based management of severe malnutrition (children) | 0% | - | - | - | 0 | 0 | 0 | 0 | 0 | 0 |
| 69 | Nutrition | Iron fortification | 60% | 24,950,160 | 50,987 | $1,172,204 | 0 | 0 | 0 | 0 | 0 | 0 |
| 70 | Nutrition | Management of severe malnutrition (children) - inpatient | 0% | - | - | - | 0 | 0 | 0 | 0 | 0 | 0 |
| 71 | Nutrition | Provision of supplementary food and nutrition counselling with growth monitoring | 0% | - | - | - | 0 | 0 | 0 | 0 | 0 | 0 |
| 72 | Nutrition | Vitamin A supplementation in infants and children 6-59 months | 0% | - | - | - | 0 | 0 | 0 | 0 | 0 | 0 |
| 73 | Nutrition | Vitamin-A fortification (sugar) and Zinc fortification (wheat) | 95% | 6,772,835 | 89,313 | $146,862 | 0 | 0 | 0 | 0 | 0 | 0 |
| 74 | Nutrition | Zinc supplementation | 0% | - | - | - | 0 | 0 | 0 | 0 | 0 | 0 |
| 75 | RMNCH | Active management of the 3rd stage of labour | 55% | 1,067,468 | 7,792,514 | $244,847 | 15 | 307 | 0 | 0 | 0 | 0 |
| 76 | RMNCH | Antenatal corticosteroids for preterm labour | 20% | 19,401 | 24,445 | $65,783 | 0 | 0 | 0 | 0 | 0 | 0 |
| 77 | RMNCH | Antibiotics for pPRoM | 55% | 49,815 | 39,354 | $44,512 | 0 | 1 | 1 | 0 | 0 | 0 |
| 78 | RMNCH | Basic ANC | 100% | 2,573,730 | 207,571 | $100,146,870 | 110 | 2959 | 0 | 0 | 0 | 0 |
| 79 | RMNCH | Cervical cancer screening | 0% | - | - | - | 0 | 0 | 0 | 0 | 0 | 0 |
| 80 | RMNCH | Cesearian section with indication | 92% | 12,123 | 116,825 | $762,583 | 8 | 23 | 2 | 0 | 0 | 0 |
| 81 | RMNCH | Cesearian Section with indication (with complication) | 92% | 2,139 | 58,168 | $231,719 | 1 | 4 | 0 | 0 | 0 | 0 |
| 82 | RMNCH | Chlorhexidine | 20% | 380,028 | 448,433 | $159,176 | 3 | 178 | 6 | 6 | 0 | 0 |
| 83 | RMNCH | Clean practices and immediate essential newborn care (in facility) | 76% | 1,470,512 | 421,613 | $2,009,097 | 2 | 56 | 0 | 0 | 0 | 0 |
| 84 | RMNCH | Condoms | 0% | - | - | - | 0 | 0 | 0 | 0 | 0 | 0 |
| 85 | RMNCH | Daily iron and folic acid supplementation (pregnant women) | 83% | 2,127,617 | 404,247 | $3,216,957 | 30 | 612 | 0 | 0 | 0 | 0 |
| 86 | RMNCH | Ectopic case management | 0% | - | - | - | 0 | 0 | 0 | 0 | 0 | 0 |
| 87 | RMNCH | Female sterilization | 73% | 256,592 | 94,939 | $1,270,129 | 514 | 295 | 18 | 0 | 0 | 0 |
| 88 | RMNCH | Fistula | 26% | 2,638 | 18,518 | $128,476 | 0 | 1 | 0 | 0 | 0 | 0 |
| 89 | RMNCH | Hypertensive disorder case management | 0% | - | - | - | 0 | 0 | 0 | 0 | 0 | 0 |
| 90 | RMNCH | Implant | 73% | 51,318 | 18,988 | $1,775,614 | 0 | 20 | 0 | 0 | 0 | 0 |
| 91 | RMNCH | Induction of labour (beyond 41 weeks) | 44% | 42,699 | 312,128 | $149 | 15 | 79 | 0 | 1 | 0 | 0 |
| 92 | RMNCH | Injectable | 73% | 2,309,325 | 854,450 | $12,516,540 | 0 | 885 | 0 | 0 | 0 | 0 |
| 93 | RMNCH | IUD | 73% | 205,273 | 75,951 | $340,754 | 0 | 79 | 0 | 0 | 0 | 0 |
| 94 | RMNCH | Kangaroo mother care | 60% | 228,017 | 417,271 | $0 | 0 | 5 | 0 | 0 | 0 | 0 |
| 95 | RMNCH | Labour and delivery management | 10% | 198,966 | 22,497 | $447,959 | 0 | 517 | 6 | 0 | 0 | 0 |
| 96 | RMNCH | Male sterilization | 73% | 234,680 | 86,832 | $1,966,617 | 470 | 270 | 16 | 0 | 0 | 0 |
| 97 | RMNCH | Management of eclampsia | 0% | - | - | - | 0 | 0 | 0 | 0 | 0 | 0 |
| 98 | RMNCH | Management of obstructed labour | 100% | 194,009 | 423,060 | $3,982,346 | 83 | 223 | 5 | 0 | 0 | 0 |
| 99 | RMNCH | Management of pre-eclampsia | 0% | - | - | - | 0 | 0 | 0 | 0 | 0 | 0 |
| 100 | RMNCH | Maternal Sepsis case management | 0% | - | - | - | 0 | 0 | 0 | 0 | 0 | 0 |
| 101 | RMNCH | Neonatal resuscitation (institutional) | 73% | 139,398 | 183,921 | $52,722 | 1 | 9 | 0 | 0 | 0 | 0 |
| 102 | RMNCH | Newborn sepsis - Full supportive care | 0% | - | - | - | 0 | 0 | 0 | 0 | 0 | 0 |
| 103 | RMNCH | Pill | 73% | 1,796,141 | 664,572 | $10,866,656 | 0 | 34 | 25 | 0 | 0 | 0 |
| 104 | RMNCH | Post-abortion case management | 0% | - | - | - | 0 | 0 | 0 | 0 | 0 | 0 |
| 105 | RMNCH | Prenatal distribution of misoprostol (for PPH prevention) | 100% | 2,573,730 | 205,898 | $920,366 | 0 | 49 | 0 | 0 | 0 | 0 |
| 106 | RMNCH | Safe abortion services | 0% | - | - | - | 0 | 0 | 0 | 0 | 0 | 0 |
| 107 | RMNCH | Support for breastfeeding mothers | 76% | 1,470,512 | 356,292 | $0 | 2 | 34 | 0 | 0 | 0 | 0 |
| 108 | RMNCH | Syphilis detection and treatment (pregnant women) | 0% | - | - | - | 0 | 0 | 0 | 0 | 0 | 0 |
| 109 | RMNCH | Tetanus toxoid (pregnant women) | 85% | 2,187,671 | 206,438 | $479,973 | 0 | 42 | 31 | 0 | 0 | 0 |
| 110 | RMNCH | Vaginal delivery - skilled attendance | 0% | - | - | - | 0 | 0 | 0 | 0 | 0 | 0 |
| 111 | RMNCH | Vaginal Delivery - with complication | 92% | 354,429 | 202,608 | $6,581,490 | 152 | 408 | 10 | 0 | 0 | 0 |
| 112 | TB | Cotrimoxazole preventive therapy for TB HIV+ patients | 0% | - | - | - | 0 | 0 | 0 | 0 | 0 | 0 |
| 113 | TB | First line treatment of smear positive cases (95% coverage) | 33% | 15,356 | 1,109,565 | $573,109 | 2 | 4 | 2 | 0 | 0 | 0 |
| 114 | TB | Full combination DOTS (smear-positive, smear negative, extrapulmonary cases, MDR cases) | 17% | 8,215 | 646,232 | $35,947,644 | 1 | 3 | 0 | 0 | 0 | 0 |
| 115 | TB | Full DOTS (smear-positive, smear negative and Extrapulmonary cases) | 33% | 15,566 | 1,225,464 | $9,684,393 | 1 | 5 | 1 | 0 | 0 | 0 |
| 116 | TB | Isonized Preventive Therapy for children in contact with TB patients | 0% | - | - | - | 0 | 0 | 0 | 0 | 0 | 0 |
| 117 | TB | Isonized Preventive Therapy for HIV+ people | 85% | 1,173,000 | 12,718,839 | $13,219,154 | 151 | 292 | 115 | 0 | 0 | 0 |
| 118 | TB | Isonized Preventive Therapy for HIV+ pregnant women | 85% | 108,230 | 2,195,671 | $1,219,706 | 14 | 27 | 11 | 0 | 0 | 0 |
| 119 | TB | Xpert test (Full) | 0% | - | - | - | 0 | 0 | 0 | 0 | 0 | 0 |
| 120 | TB | Xpert test (targeted) | 0% | - | - | - | 0 | 0 | 0 | 0 | 0 | 0 |
| 121 | Vaccine Preventable Diseases | BCG vaccine | 0% | - | - | - | 0 | 0 | 0 | 0 | 0 | 0 |
| 122 | Vaccine Preventable Diseases | DPT-Heb-Hib / Pentavalent vaccine | 90% | 1,675,582 | 6,995,270 | $3,080,496 | 0 | 32 | 23 | 0 | 0 | 0 |
| 123 | Vaccine Preventable Diseases | HPV vaccine | 0% | - | - | - | 0 | 0 | 0 | 0 | 0 | 0 |
| 124 | Vaccine Preventable Diseases | Measles vaccine | 0% | - | - | - | 0 | 0 | 0 | 0 | 0 | 0 |
| 125 | Vaccine Preventable Diseases | Oral cholera vaccine | 0% | - | - | - | 0 | 0 | 0 | 0 | 0 | 0 |
| 126 | Vaccine Preventable Diseases | Pneumococcal vaccine | 90% | 1,675,582 | 5,864,537 | $15,562,898 | 0 | 32 | 23 | 0 | 0 | 0 |
| 127 | Vaccine Preventable Diseases | Polio vaccine | 0% | - | - | - | 0 | 0 | 0 | 0 | 0 | 0 |
| 128 | Vaccine Preventable Diseases | Rotavirus vaccine | 90% | 1,675,582 | 314,959 | $7,786,880 | 0 | 32 | 23 | 0 | 0 | 0 |

Supplementary Table 4: Health outcomes and resource use by intervention under the task-shifting scenario

| **No.** | **Program** | **Intervention** | **% of cases in need covered under optimal package** | **Total number of cases covered under the optimal package** | **DALYs averted** | **Consumables expenditure required** | **Medical/Clinical Officer** | **Nursing staff** | **Pharmaceutical staff** | **Lab staff** | **Mental health staff** | **Nutrition staff** |
| --- | --- | --- | --- | --- | --- | --- | --- | --- | --- | --- | --- | --- |
| 1 | HIV & STIs | ART (Second-Line Treatment) for adults with intensive monitoring | 0% | 0 | 0 | $0 | 0 | 0 | 0 | 0 | 0 | 0 |
| 2 | HIV & STIs | ART (Second-Line Treatment) for adults without intensive monitoring | 0% | 0 | 0 | $0 | 0 | 0 | 0 | 0 | 0 | 0 |
| 3 | HIV & STIs | ART for men | 0% | 0 | 0 | $0 | 0 | 0 | 0 | 0 | 0 | 0 |
| 4 | HIV & STIs | ART for women | 0% | 0 | 0 | $0 | 0 | 0 | 0 | 0 | 0 | 0 |
| 5 | HIV & STIs | Blood safety | 100% | 415,836 | 193,120 | $2,813,131 | 1 | 0 | 0 | 10 | 0 | 0 |
| 6 | HIV & STIs | HIV Testing Services | 0% | 0 | 0 | $0 | 0 | 0 | 0 | 0 | 0 | 0 |
| 7 | HIV & STIs | Home-based highly active retroviral therapy (HAART) | 0% | 0 | 0 | $0 | 0 | 0 | 0 | 0 | 0 | 0 |
| 8 | HIV & STIs | Interventions focused on male sex workers | 75% | 1,661 | 11,413 | $147,500 | 0 | 1 | 0 | 0 | 0 | 0 |
| 9 | HIV & STIs | Interventions focused on men who have sex with men | 75% | 18,075 | 124,175 | $2,139,638 | 0 | 12 | 0 | 0 | 0 | 0 |
| 10 | HIV & STIs | Male circumcision | 40% | 1,094,584 | 339,321 | $11,283,038 | 313 | 420 | 0 | 0 | 0 | 0 |
| 11 | HIV & STIs | Mass media | 71% | 29,524,356 | 250,311 | $0 | 0 | 0 | 0 | 0 | 0 | 0 |
| 12 | HIV & STIs | Peer education for sex workers | 71% | 31,453 | 928,931 | $0 | 0 | 0 | 0 | 0 | 0 | 0 |
| 13 | HIV & STIs | PMTCT | 99% | 73,586 | 631,366 | $1,615,645 | 6 | 32 | 0 | 0 | 0 | 0 |
| 14 | HIV & STIs | Pre-exposure prophylaxis for high-risk serodiscordant couples | 0% | 0 | 0 | $0 | 0 | 0 | 0 | 0 | 0 | 0 |
| 15 | HIV & STIs | Pre-exposure prophylaxis for pregnant and breastfeeding women | 0% | 0 | 0 | $0 | 0 | 0 | 0 | 0 | 0 | 0 |
| 16 | HIV & STIs | Screen HIV+ cases for TB | 60% | 1,005,838 | 1,028,700 | $12,037,082 | 173 | 145 | 0 | 0 | 0 | 0 |
| 17 | HIV & STIs | Treatment of chlamydia | 70% | 15389697.09 | 389372.0965 | $54,760,419 | 1100 | 3893 | 0 | 0 | 0 | 0 |
| 18 | HIV & STIs | Treatment of gonorrhea | 70% | 15389697.09 | 389372.0965 | $12,585,074 | 1100 | 3893 | 0 | 0 | 0 | 0 |
| 19 | HIV & STIs | Treatment of PID (Pelvic Inflammatory Disease) | 70% | 3885898.515 | 389372.0965 | $21,159,039 | 278 | 983 | 0 | 0 | 0 | 0 |
| 20 | HIV & STIs | Treatment of trichomoniasis | 70% | 15389697.09 | 389372.0965 | $6,752,966 | 1100 | 3893 | 0 | 0 | 0 | 0 |
| 21 | HIV & STIs | Youth focused interventions - In-school | 0% | 0 | 0 | $0 | 0 | 0 | 0 | 0 | 0 | 0 |
| 22 | IMCI | Antibiotics for treatment of dysentery | 0% | 0 | 0 | $0 | 0 | 0 | 0 | 0 | 0 | 0 |
| 23 | IMCI | ORS and Zinc for diarrhea | 0% | 0 | 0 | $0 | 0 | 0 | 0 | 0 | 0 | 0 |
| 24 | IMCI | Pneumonia treatment (children) | 0% | 0 | 0 | $0 | 0 | 0 | 0 | 0 | 0 | 0 |
| 25 | IMCI | Treatment of severe diarrhea | 100% | 199,339 | 117,610 | $416,064 | 31 | 27 | 4 | 0 | 0 | 0 |
| 26 | Malaria | Case management with artemisinin based combination therapy (80% coverage) | 34% | 8,597,325 | 134,577 | $21,863,282 | 1168 | 824 | 0 | 0 | 0 | 0 |
| 27 | Malaria | Complicated (children, injectable artesunate) | 100% | 27,947 | 19,563 | $360,158 | 4 | 5 | 0 | 0 | 0 | 0 |
| 28 | Malaria | Home management of fevers using antimalarial (artesunate- amodiaquine AAQ) - under 5 | 87% | 2,101,921 | 6,499,956 | $5,345,255 | 0 | 282 | 47 | 0 | 0 | 0 |
| 29 | Malaria | Indoor residual spray and LLINs | 70% | 1,996,803 | 1,522,962 | $18,470,427 | 0 | 38 | 0 | 0 | 0 | 0 |
| 30 | Malaria | Intermittent preventive treatment in infants (IPTi) using 3 days of amodiaquine-artesunate (AQ3-AS3) against clinical malaria; at 2, 3, and 9 months. | 41% | 3,301,868 | 521,695 | $4,107,524 | 519 | 443 | 74 | 0 | 0 | 0 |
| 31 | Malaria | IPT (pregnant women) | 0% | 0 | 0 | $0 | 0 | 0 | 0 | 0 | 0 | 0 |
| 32 | Malaria | ITN distribution to pregnant women | 0% | 0 | 0 | $0 | 0 | 0 | 0 | 0 | 0 | 0 |
| 33 | Mental Health | Anti-epileptic medication | 10% | 19,629 | 6,021 | $156,164 | 0 | 0 | 0 | 0 | 4 | 0 |
| 34 | Mental Health | Basic psychosocial support, advice, and follow-up | 0% | 0 | 0 | $0 | 0 | 0 | 0 | 0 | 0 | 0 |
| 35 | Mental Health | Treatment of acute psychotic disorders | 0% | 0 | 0 | $0 | 0 | 0 | 0 | 0 | 0 | 0 |
| 36 | Mental Health | Treatment of bipolar disorder | 0% | 0 | 0 | $0 | 0 | 0 | 0 | 0 | 0 | 0 |
| 37 | Mental Health | Treatment of depression | 0% | 0 | 0 | $0 | 0 | 0 | 0 | 0 | 0 | 0 |
| 38 | Mental Health | Treatment of schizophrenia | 0% | 0 | 0 | $0 | 0 | 0 | 0 | 0 | 0 | 0 |
| 39 | NCDs | Amputation | 90% | 112.27572 | 2,387 | $87,375 | 1 | 0 | 0 | 0 | 0 | 0 |
| 40 | NCDs | Asthma: Inhaled short acting beta agonist for intermittent asthma | 0% | 0 | 0 | $0 | 0 | 0 | 0 | 0 | 0 | 0 |
| 41 | NCDs | Asthma: Low dose inhaled beclometasone + SABA | 0% | 0 | 0 | $0 | 0 | 0 | 0 | 0 | 0 | 0 |
| 42 | NCDs | Breast Cancer (clinical examination + treatment) | 0% | 0 | 0 | $0 | 0 | 0 | 0 | 0 | 0 | 0 |
| 43 | NCDs | Breast Cancer (first line) | 0% | 0 | 0 | $0 | 0 | 0 | 0 | 0 | 0 | 0 |
| 44 | NCDs | Breast Cancer (mammography + treatment) | 0% | 0 | 0 | $0 | 0 | 0 | 0 | 0 | 0 | 0 |
| 45 | NCDs | Cervical cancer (first line) | 0% | 0 | 0 | $0 | 0 | 0 | 0 | 0 | 0 | 0 |
| 46 | NCDs | Colorectoral cancer (screening + treatment) | 0% | 0 | 0 | $0 | 0 | 0 | 0 | 0 | 0 | 0 |
| 47 | NCDs | Colorectoral cancer (treatment) | 0% | 0 | 0 | $0 | 0 | 0 | 0 | 0 | 0 | 0 |
| 48 | NCDs | community-based management of hypertension | 0% | 0 | 0 | $0 | 0 | 0 | 0 | 0 | 0 | 0 |
| 49 | NCDs | COPD - Inhaled salbutamol | 0% | 0 | 0 | $0 | 0 | 0 | 0 | 0 | 0 | 0 |
| 50 | NCDs | COPD - oxygen therapy and drugs | 0% | 0 | 0 | $0 | 0 | 0 | 0 | 0 | 0 | 0 |
| 51 | NCDs | COPD - treatment of severe exacerbations | 0% | 0 | 0 | $0 | 0 | 0 | 0 | 0 | 0 | 0 |
| 52 | NCDs | Elective inguinal hernia repair | 50% | 215.818884 | 1,322 | $19,754 | 1 | 1 | 0 | 0 | 0 | 0 |
| 53 | NCDs | Emergency inguinal hernia repair | 90% | 654.9417 | 23,469 | $59,947 | 1 | 0 | 0 | 0 | 0 | 0 |
| 54 | NCDs | GIT, Intestine cancer | 0% | 0 | 0 | $0 | 0 | 0 | 0 | 0 | 0 | 0 |
| 55 | NCDs | Prevention and treatment of cardiovascular disease | 0% | 0 | 0 | $0 | 0 | 0 | 0 | 0 | 0 | 0 |
| 56 | NCDs | Prevention of cardiovascular disease | 0% | 0 | 0 | $0 | 0 | 0 | 0 | 0 | 0 | 0 |
| 57 | NCDs | Retinopathy Screening and photocoagulation for diabetics | 0% | 0 | 0 | $0 | 0 | 0 | 0 | 0 | 0 | 0 |
| 58 | NCDs | Substance use disorder - alcohol | 0% | 0 | 0 | $0 | 0 | 0 | 0 | 0 | 0 | 0 |
| 59 | NCDs | Testing of pre-cancerous cells (vinegar) | 1% | 11,172 | 673.5535048 | $33,407 | 2 | 1 | 0 | 0 | 0 | 0 |
| 60 | NCDs | Treatment of injuries (Fracture and dislocation - fixation) | 0% | 0 | 0 | $0 | 0 | 0 | 0 | 0 | 0 | 0 |
| 61 | NCDs | Treatment of injuries (Fracture and dislocation - reduction) | 90% | 374,252 | 603,164 | $1,946,112 | 300 | 179 | 0 | 0 | 0 | 0 |
| 62 | NTDs | Schistosomiasis Mass drug administration (adults) | 80% | 5,107,568 | 620,391 | $1,833,701 | 0 | 98 | 0 | 0 | 0 | 0 |
| 63 | NTDs | Trachoma mass drug administration | 80% | 10,303,760 | 208,421 | $5,467,939 | 0 | 197 | 0 | 0 | 0 | 0 |
| 64 | NTDs | Trachoma Trichiasis cases surgey | 0% | 0 | 0 | $0 | 0 | 0 | 0 | 0 | 0 | 0 |
| 65 | Nutrition | Calcium Supplementation | 0% | 0 | 0 | $0 | 0 | 0 | 0 | 0 | 0 | 0 |
| 66 | Nutrition | Community management of nutrition in under-5 - micronutrient powder | 0% | 0 | 0 | $0 | 0 | 0 | 0 | 0 | 0 | 0 |
| 67 | Nutrition | Community-based management of moderate acute malnutrition (children) | 7% | 113,963 | 444,456 | $5,010,851 | 0 | 0 | 2 | 0 | 0 | 46 |
| 68 | Nutrition | Community-based management of severe malnutrition (children) | 0% | 0 | 0 | $0 | 0 | 0 | 0 | 0 | 0 | 0 |
| 69 | Nutrition | Iron fortification | 60% | 24,950,160 | 50,987 | $1,172,204 | 0 | 0 | 0 | 0 | 0 | 0 |
| 70 | Nutrition | Management of severe malnutrition (children) - inpatient | 0% | 0 | 0 | $0 | 0 | 0 | 0 | 0 | 0 | 0 |
| 71 | Nutrition | Provision of supplementary food and nutrition counselling with growth monitoring | 0% | 0 | 0 | $0 | 0 | 0 | 0 | 0 | 0 | 0 |
| 72 | Nutrition | Vitamin A supplementation in infants and children 6-59 months | 0% | 0 | 0 | $0 | 0 | 0 | 0 | 0 | 0 | 0 |
| 73 | Nutrition | Vitamin-A fortification (sugar) and Zinc fortification (wheat) | 95% | 6,772,835 | 89,313 | $146,862 | 0 | 0 | 0 | 0 | 0 | 0 |
| 74 | Nutrition | Zinc supplementation | 0% | 0 | 0 | $0 | 0 | 0 | 0 | 0 | 0 | 0 |
| 75 | RMNCH | Active management of the 3rd stage of labour | 55% | 1,067,468 | 7,792,514 | $244,847 | 15 | 307 | 0 | 0 | 0 | 0 |
| 76 | RMNCH | Antenatal corticosteroids for preterm labour | 20% | 19,401 | 24,445 | $65,783 | 0 | 0 | 0 | 0 | 0 | 0 |
| 77 | RMNCH | Antibiotics for pPRoM | 55% | 49,815 | 39,354 | $44,512 | 0 | 3 | 0 | 0 | 0 | 0 |
| 78 | RMNCH | Basic ANC | 4% | 94,854 | 7,650 | $3,690,864 | 4 | 109 | 0 | 0 | 0 | 0 |
| 79 | RMNCH | Cervical cancer screening | 0% | 0 | 0 | $0 | 0 | 0 | 0 | 0 | 0 | 0 |
| 80 | RMNCH | Cesearian section with indication | 92% | 12,123 | 116,825 | $762,583 | 8 | 23 | 2 | 0 | 0 | 0 |
| 81 | RMNCH | Cesearian Section with indication (with complication) | 92% | 2,139 | 58,168 | $231,719 | 1 | 4 | 0 | 0 | 0 | 0 |
| 82 | RMNCH | Chlorhexidine | 20% | 380,028 | 448,433 | $159,176 | 3 | 178 | 6 | 6 | 0 | 0 |
| 83 | RMNCH | Clean practices and immediate essential newborn care (in facility) | 76% | 1,470,512 | 421,613 | $2,009,097 | 2 | 56 | 0 | 0 | 0 | 0 |
| 84 | RMNCH | Condoms | 0% | 0 | 0 | $0 | 0 | 0 | 0 | 0 | 0 | 0 |
| 85 | RMNCH | Daily iron and folic acid supplementation (pregnant women) | 83% | 2,127,617 | 404,247 | $3,216,957 | 30 | 612 | 0 | 0 | 0 | 0 |
| 86 | RMNCH | Ectopic case management | 0% | 0 | 0 | $0 | 0 | 0 | 0 | 0 | 0 | 0 |
| 87 | RMNCH | Female sterilization | 73% | 256,592 | 94,939 | $1,270,129 | 514 | 295 | 18 | 0 | 0 | 0 |
| 88 | RMNCH | Fistula | 26% | 2,638 | 18,518 | $128,476 | 0 | 1 | 0 | 0 | 0 | 0 |
| 89 | RMNCH | Hypertensive disorder case management | 0% | 0 | 0 | $0 | 0 | 0 | 0 | 0 | 0 | 0 |
| 90 | RMNCH | Implant | 73% | 51,318 | 18,988 | $1,775,614 | 0 | 20 | 0 | 0 | 0 | 0 |
| 91 | RMNCH | Induction of labour (beyond 41 weeks) | 44% | 42,699 | 312,128 | $149 | 15 | 79 | 0 | 1 | 0 | 0 |
| 92 | RMNCH | Injectable | 73% | 2,309,325 | 854,450 | $12,516,540 | 0 | 885 | 0 | 0 | 0 | 0 |
| 93 | RMNCH | IUD | 73% | 205,273 | 75,951 | $340,754 | 0 | 79 | 0 | 0 | 0 | 0 |
| 94 | RMNCH | Kangaroo mother care | 60% | 228,017 | 417,271 | $0 | 0 | 5 | 0 | 0 | 0 | 0 |
| 95 | RMNCH | Labour and delivery management | 43% | 837,657 | 94,713 | $1,885,928 | 0 | 2211 | 0 | 0 | 0 | 0 |
| 96 | RMNCH | Male sterilization | 73% | 234,680 | 86,832 | $1,966,617 | 470 | 270 | 16 | 0 | 0 | 0 |
| 97 | RMNCH | Management of eclampsia | 0% | 0 | 0 | $0 | 0 | 0 | 0 | 0 | 0 | 0 |
| 98 | RMNCH | Management of obstructed labour | 100% | 194,009 | 423,060 | $3,982,346 | 83 | 223 | 5 | 0 | 0 | 0 |
| 99 | RMNCH | Management of pre-eclampsia | 0% | 0 | 0 | $0 | 0 | 0 | 0 | 0 | 0 | 0 |
| 100 | RMNCH | Maternal Sepsis case management | 0% | 0 | 0 | $0 | 0 | 0 | 0 | 0 | 0 | 0 |
| 101 | RMNCH | Neonatal resuscitation (institutional) | 73% | 139,398 | 183,921 | $52,722 | 1 | 9 | 0 | 0 | 0 | 0 |
| 102 | RMNCH | Newborn sepsis - Full supportive care | 70% | 1330098.506 | 98714.05263 | $2,605,176 | 399 | 762 | 0 | 53 | 0 | 0 |
| 103 | RMNCH | Pill | 73% | 1,796,141 | 664,572 | $10,866,656 | 0 | 34 | 25 | 0 | 0 | 0 |
| 104 | RMNCH | Post-abortion case management | 0% | 0 | 0 | $0 | 0 | 0 | 0 | 0 | 0 | 0 |
| 105 | RMNCH | Prenatal distribution of misoprostol (for PPH prevention) | 100% | 2,573,730 | 205,898 | $920,366 | 0 | 49 | 0 | 0 | 0 | 0 |
| 106 | RMNCH | Safe abortion services | 0% | 0 | 0 | $0 | 0 | 0 | 0 | 0 | 0 | 0 |
| 107 | RMNCH | Support for breastfeeding mothers | 76% | 1,470,512 | 356,292 | $0 | 2 | 34 | 0 | 0 | 0 | 0 |
| 108 | RMNCH | Syphilis detection and treatment (pregnant women) | 59% | 1526262.295 | 151087.3051 | $733,344 | 109 | 339 | 35 | 0 | 0 | 0 |
| 109 | RMNCH | Tetanus toxoid (pregnant women) | 85% | 2,187,671 | 206,438 | $479,973 | 0 | 84 | 0 | 0 | 0 | 0 |
| 110 | RMNCH | Vaginal delivery - skilled attendance | 76% | 1470511.968 | 166269.8262 | $3,310,758 | 0 | 3881 | 0 | 0 | 0 | 0 |
| 111 | RMNCH | Vaginal Delivery - with complication | 92% | 354,429 | 202,608 | $6,581,490 | 152 | 408 | 10 | 0 | 0 | 0 |
| 112 | TB | Cotrimoxazole preventive therapy for TB HIV+ patients | 0% | 0 | 0 | $0 | 0 | 0 | 0 | 0 | 0 | 0 |
| 113 | TB | First line treatment of smear positive cases (95% coverage) | 17% | 7,664 | 553,777 | $286,035 | 1 | 2 | 1 | 0 | 0 | 0 |
| 114 | TB | Full combination DOTS (smear-positive, smear negative, extrapulmonary cases, MDR cases) | 33% | 15,906 | 1,251,318 | $69,606,507 | 1 | 5 | 1 | 0 | 0 | 0 |
| 115 | TB | Full DOTS (smear-positive, smear negative and Extrapulmonary cases) | 33% | 15,566 | 1,225,464 | $9,684,393 | 1 | 5 | 1 | 0 | 0 | 0 |
| 116 | TB | Isonized Preventive Therapy for children in contact with TB patients | 85% | 39131.53823 | 18420.52739 | $86,089 | 5 | 10 | 4 | 0 | 0 | 0 |
| 117 | TB | Isonized Preventive Therapy for HIV+ people | 85% | 1,173,000 | 12,718,839 | $13,219,154 | 151 | 292 | 115 | 0 | 0 | 0 |
| 118 | TB | Isonized Preventive Therapy for HIV+ pregnant women | 85% | 108,230 | 2,195,671 | $1,219,706 | 14 | 27 | 11 | 0 | 0 | 0 |
| 119 | TB | Xpert test (Full) | 0% | 0 | 0 | $0 | 0 | 0 | 0 | 0 | 0 | 0 |
| 120 | TB | Xpert test (targeted) | 0% | 0 | 0 | $0 | 0 | 0 | 0 | 0 | 0 | 0 |
| 121 | Vaccine Preventable Diseases | BCG vaccine | 90% | 1675582.091 | 192042.225 | $949,966 | 0 | 32 | 23 | 0 | 0 | 0 |
| 122 | Vaccine Preventable Diseases | DPT-Heb-Hib / Pentavalent vaccine | 90% | 1,675,582 | 6,995,270 | $3,080,496 | 0 | 32 | 23 | 0 | 0 | 0 |
| 123 | Vaccine Preventable Diseases | HPV vaccine | 0% | 0 | 0 | $0 | 0 | 0 | 0 | 0 | 0 | 0 |
| 124 | Vaccine Preventable Diseases | Measles vaccine | 90% | 1675582.091 | 9464.488288 | $1,184,742 | 0 | 32 | 23 | 0 | 0 | 0 |
| 125 | Vaccine Preventable Diseases | Oral cholera vaccine | 0% | 0 | 0 | $0 | 0 | 0 | 0 | 0 | 0 | 0 |
| 126 | Vaccine Preventable Diseases | Pneumococcal vaccine | 90% | 1,675,582 | 5,864,537 | $15,562,898 | 0 | 32 | 23 | 0 | 0 | 0 |
| 127 | Vaccine Preventable Diseases | Polio vaccine | 0% | 0 | 0 | $0 | 0 | 0 | 0 | 0 | 0 | 0 |
| 128 | Vaccine Preventable Diseases | Rotavirus vaccine | 90% | 1,675,582 | 314,959 | $7,786,880 | 0 | 64 | 0 | 0 | 0 | 0 |

Supplementary table 5: List of interventions treated as substitutes

| **Group 1** | 1. First line treatment of smear positive cases, 2. Full DOTS (smear-positive, smear negative and Extrapulmonary cases), 3. Full combination DOTS (smear-positive, smear negative, extrapulmonary cases, MDR cases) |
| --- | --- |
| **Group 2** | 1. Prevention of cardiovascular disease, 2. Prevention and treatment of cardiovascular disease |
| **Group 3** | 1. Xpert test (Full), 2. Xpert test (targeted) |
| **Group 4** | 1. Colorectoral cancer (screening + treatment), 2. Colorectoral cancer (treatment) |
| **Group 5** | 1. ART (Second-Line Treatment) for adults without intensive monitoring, 2. ART (Second-Line Treatment) for adults with intensive monitoring |

Supplementary Table 6: Monthly salaries used in marginal value calculation

| **Health worker cadre** | **Monthly salary (2019 USD)** |
| --- | --- |
| Doctor/Medical officer | 567 |
| Nurse | 166 |
| Pharmacist | 230 |
| Laboratory staff | 230 |
| Mental health officer | 230 |
| Nutrition officer | 166 |

Supplementary table 7: Summary of results from constrained optimization under scenarios with different constraints

| **Scenario** | **Base scenario** | **Task-shifting scenario** | **Only consumables budget constraint scenario** | **Only consumables budget and feasible coverage constraint scenario** |
| --- | --- | --- | --- | --- |
| Number of interventions with positive NHB | 83 | 83 | 83 | 83 |
| Number of interventions in the optimal package | 58 | 68 | 62 | 72 |
| Total DALYs averted | 59,303,843.0 | 64,734,224.0 | 86,929,541.0 | 65,213,603.0 |
| Highest ICER in the HBP | 122.2 | 122.2 | 122.2 | 143.5 |
| % of consumables budget required | 100% | 100% | 100% | 100% |
| % of Medical/Clinical officer capacity required | 78% | 100% | 126% | 133% |
| % of Nursing staff capacity required | 44% | 100% | 115% | 102% |
| % of pharmaceutical staff capacity required | 100% | 100% | 685% | 722% |
| % of Lab staff capacity required | 0% | 2% | 3% | 2% |
| % of Mental health staff capacity required | 1% | 1% | 0% | 1% |
| % of Nutrition staff capacity required | 100% | 100% | 1689% | 1089% |
